# Dissecting the genetic and proteomic risk factors for delirium

**DOI:** 10.1101/2024.10.11.24315324

**Authors:** Vasilis Raptis, Youngjune Bhak, Timothy I Cannings, Alasdair M. J. MacLullich, Albert Tenesa

**Affiliations:** The Roslin Institute, Royal (Dick) School of Veterinary Studies, The University of Edinburgh, Easter Bush Campus, Midlothian, UK; Advanced Care Research Centre, School of Engineering, College of Science and Engineering, The University of Edinburgh, Edinburgh, UK; School of Mathematics and Maxwell Institute for Mathematical Sciences, University of Edinburgh, UK; Edinburgh Delirium Research Group, Ageing and Health, Usher Institute, University of Edinburgh, Edinburgh, Scotland, UK; Institute of Genetics and Cancer, University of Edinburgh, Western General Hospital, Edinburgh, UK

## Abstract

Delirium is an acute change in cognition, common in hospitalised older adults, and associated with high healthcare and human cost. In this work we shed light on the currently poorly understood genetic and proteomic background of delirium. We conducted the largest to date multi-ancestry analysis of genetic variants associated with delirium (1,059,130 individuals, 11,931 cases), yielding the *Apolipoprotein E* (*APOE*) gene as a strong risk factor with possible population and age-varying effects. *APOE* genetic effect on delirium remained significant after adjusting for dementia and Alzheimer disease (AD) and in dementia-free cohorts. A multi-trait analysis of delirium with AD identified 5 delirium genetic risk loci. Investigation of plasma proteins associated with up to 16-years incident delirium (32,652 individuals, 541 cases) revealed known and novel protein biomarkers, implicating brain vulnerability, inflammation and immune response processes. Incorporating proteomic and genetic evidence via mendelian randomisation, colocalisation and druggability analyses, we indicate putatively useful drug target proteins for delirium. Integrating proteins and *APOE* genetic risk with demographics significantly improved incident delirium prediction compared to demographics alone. Our results pave the way to better understanding delirium’s aetiology and guide further research on clinically relevant biomarkers.

---

Delirium is a complex neurocognitive condition affecting nearly 25% of hospitalised older adults^1^. It has a rapid onset, usually lasting from hours to days, and is characterised by an acute and often reversible disturbance of attention, cognitive ability and awareness^2^. It is triggered mainly by acute illness, surgery or injury. Delirium is strongly associated with multiple adverse outcomes, including increased mortality, prolonged hospitalisation^3^, and increased healthcare costs, with one study estimating an annual cost of $182 billion dollars per year to European healthcare systems^4^.

Delirium has a complex bidirectional relationship with dementia, in that people with dementia are more likely to experience delirium in the context of an acute trigger, and delirium episodes are strongly associated with future dementia risk^3,5^. There is some overlap in the clinical features of these conditions, notably with respect to cognitive dysfunction, perceptual disturbances and sleep-wake cycle changes^6^. However, the course of delirium is markedly different with respect to onset and duration. The pathophysiology of delirium is complex and likely involves multiple potential mechanisms^3^. Animal model and human biomarker studies implicate systemic inflammation triggering neuroinflammation, disruption of the blood-brain barrier, and neuronal injury in delirium^3,7–9^. Aspects of these mechanisms are also present in dementia, leading to the proposal that there may be shared and potentially bidirectional pathology between delirium and dementia, such as neuroinflammation leading to neurodegeneration^9,10^.

Despite its high healthcare burden, however, current understanding of genetic and biological mechanisms underlying delirium’s pathophysiology is still limited, hindering personalised medicine efforts to predict, prevent and treat the condition. Given delirium’s increasing presence in the global ageing population^6^, alleviating its human and economic cost^11^ through personalised medicine is all the more important.

Previous studies on the genetic determinants of delirium have been small in scale and inconclusive^3,6,12^, largely focusing on a single or small sets of candidate genes^13–22^. The apolipoprotein E gene (*APOE*), specifically its ε4 haplotype, is the most intensively studied gene^15,17,19–21^, though clear conclusions on its relationship with delirium remain lacking^6,23^. Advances in genome-wide association studies (GWAS) over the last decades^24^ have allowed researchers to search across the full spectrum of the human genome for genetic risk factors involved in neurocognitive disorders^25–27^, offering invaluable new insights into disease mechanisms. However, research on delirium using these methods has lagged behind other neurocognitive disorders, with only a few, relatively underpowered delirium GWAS conducted to date^28–30^. Moreover, those studies did not address comprehensively the biological implications of the potential gene-disease associations and were conducted primarily on individuals of European descent.

Research on protein biomarkers for delirium has received increased attention recently, identifying potential blood plasma, serum and cerebrospinal fluid (CSF) canditates^6,31^. Those biomarkers include inflammatory proteins, such as interleukin-6 (IL-6) and C-reactive protein (CRP)^6,31,32^, Alzheimer disease pathology markers^6^, and markers of neuronal injury, such as the neurofilament light chain (NEFL)^9^ and glial fibrillary acidic protein (GFAP)^8^. Previous studies however are limited in terms of sample size and, often, the range of proteins tested.

In the current study we aim to upscale the efforts in identifying genetic and proteomic determinants of delirium risk. To achieve this, we: (1) conducted the largest so far meta-analysis of delirium GWAS datasets, the first to include individuals from diverse ancestries; (2) tested for plasma proteome signatures of incident delirium for up to 16 years of follow-up in UK Biobank (UKB)^33^ and triangulated the results using genetically supported evidence from mendelian randomisation, colocalisation and druggability analyses; (3) conducted a multi-trait meta-analysis between delirium and Alzheimer disease, leveraging the shared genetic basis of the two conditions^6^ and boosting the power to detect genetic associations for delirium^34^.

## Results

### *APOE* gene as delirium genetic risk factor

To identify genetic variants associated with delirium, a multi-ancestry genome-wide association meta-analysis (GWAMA) was conducted on eight sub-cohorts (Supplementary Table 1) from four global ancestries: European (EUR, *n*_*cases*_ = 7,988, *n*_*controls*_ = 549,568, 52.6% of total sample size), Finnish (FIN, *n*_*cases*_ = 3,371, *n*_*controls*_ = 388,560, 37%), African (AFR, *n*_*cases*_ = 348; *n*_*controls*_ = 59,780, 5.7%), South Asian (SAS, *n*_*cases*_ = 107; *n*_*controls*_ = 9,356, 0.9%) and admixed American / Hispanic (AMR, *n*_*cases*_ = 117; *n*_*controls*_ = 39,977, 3.8%). Age distributions for the UK Biobank (UKB) and All of Us Research Program (AoU) cohorts are presented in Supplementary Figure 1. In total, the GWAMA comprised of up to 1,059,130 individuals and 11,931 delirium cases, yielding results for 24,951,028 single nucleotide polymorphism (SNP) genetic variants. The overall SNP heritability (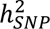) of delirium was estimated on the observed scale as 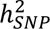 = 0.0055 (*standard error*: 9×10^−4^; *p* = 2.5×10^−9^), significantly different from zero, and as 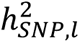 = 0.029 on the liability scale.

Variants at the *Apolipoprotein E* (*APOE*) gene and within its close genomic region on chromosome 19 (Figure 1) were significantly associated with delirium. The lead variant rs429358 (T>C, odds ratio [95% Confidence Intervals]: 1.60 [1.55 – 1.65]; p = 9.7x10^-177^) is an *APOE* missense variant, which together with the rs7412 C>T variant forms the *APOE*-ε4 haplotype (the rs429358-C and rs7412-C alleles), an established risk factor for Alzheimer disease (AD)^35^. rs7412 also significantly associated with delirium in our GWAMA (C>T, OR [95% CI]: 0.84 [0.79 – 0.88]; p = 1.8x10^-11^).

**Figure 1:**
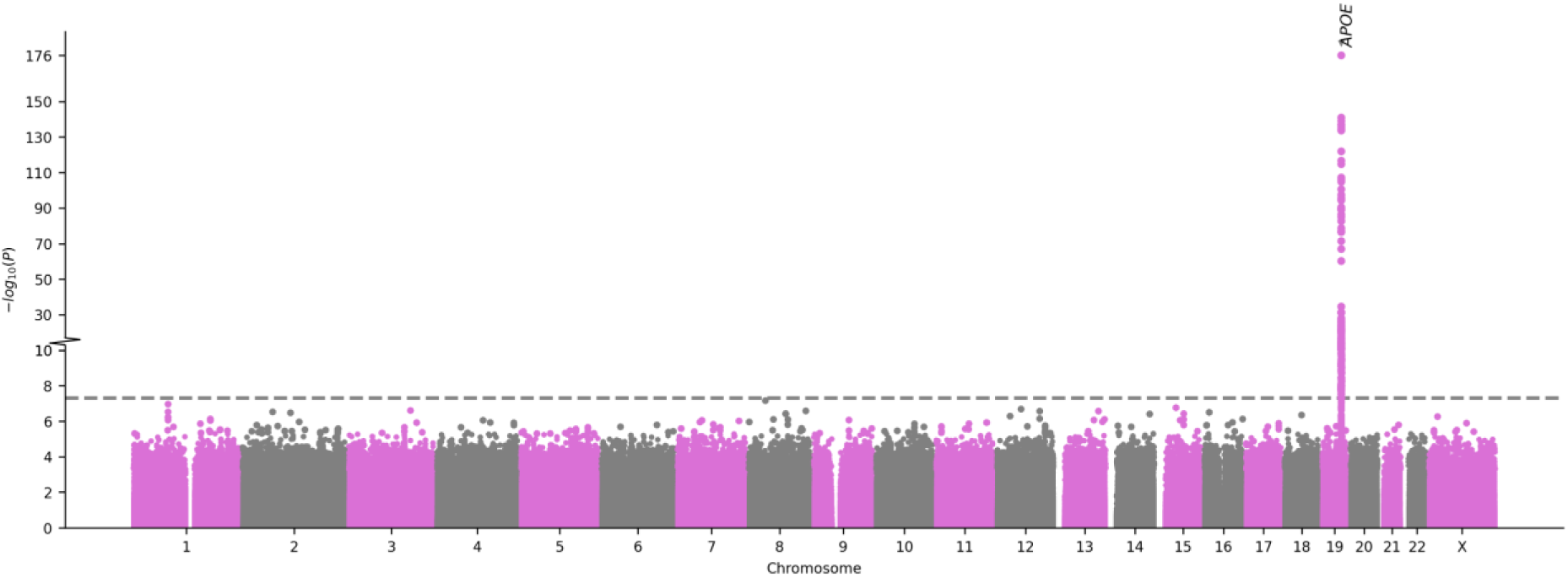
Manhattan plot of the delirium multi-ancestry GWAMA. Each point represents a genetic variant. The x-axis denotes the variant’s genomic position and the y-axis the two-sided meta-analysis association p-value. The grey dashed line denotes the genome-wide significance p-value threshold 5x10^-8^. The gene in which the lead significant variant is located is annotated. GWAMA: genome-wide association meta-analysis; APOE: Apolipoprotein E gene. The plot was created using the GWASLab python package.

The lead variant rs429358 showed population specific genetic effects for delirium in the contributing sub-cohorts (Figure 2). Significant associations were observed in all, except European (Michigan Genomics Initiative (MGI), p = 0.27) and admixed American (AoU AMR, p = 0.12) populations from the USA, with USA-based populations generally showing smaller effect sizes.

**Figure 2:**
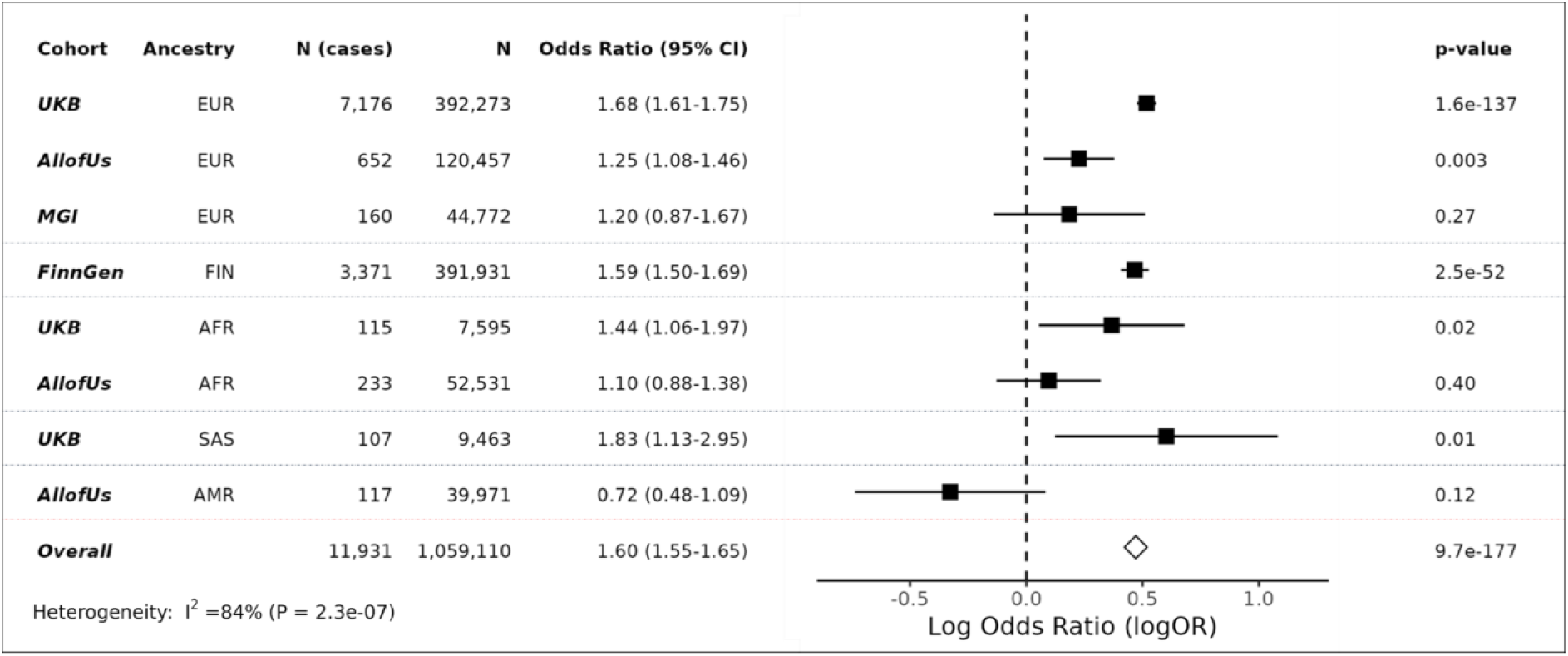
Associations between the GWAMA lead variant and delirium risk for each cohort. Forrest plot showing the cohort-specific association results for the rs429358 T>C lead variant with delirium. Effect sizes for the C allele and corresponding two-sided p-values are shown for each contributing sub-cohort and the overall meta-analysis. Grey dotted lines separate results for each ancestry group. GWAMA: genome-wide association meta-analysis; UKB: UK Biobank; MGI: Michigan Genomics Initiative; N: sample size; CI: 95% confidence intervals for the odds ratio; EUR: European; FIN: Finnish; AFR: African; SAS: south Asian; AMR: admixed American).

Conditional GWAS analysis on the *APOE-*ε4 haplotype resulted in all variants on the *APOE* region losing significance (Supplementary Figure 2), suggesting that *APOE-*ε4 is the sole independent genetic risk factor in the region. On the same analysis, an intronic variant within the *ADAM32* gene on chromosome 8 gained significance (rs531178459; p = 3.3x10^-8^).

Furthermore, using a series of sensitivity analyses in UKB (see Methods and Supplementary Table 2), we tested to what extent the observed *APOE* association with delirium is driven by underlying dementia. Variants in the *APOE* region remained significant after adjusting for dementia status in the delirium GWAS (Supplementary Figure 3). Specifically, the lead variant from the unadjusted GWAMA, rs429358, showed again a strong association (OR [95% CI]: 1.20 [1.12 – 1.28]; p = 3.7x10^-15^). Variants on the *SEC14L1* gene on chromosome 17 were also significant in the dementia-adjusted analysis. rs429358 remained significant in GWAS conducted on dementia-free (OR [95% CI]: 1.27 [1.2 – 1.34]; p = 4.7x10^-18^; Supplementary Figure 4) and AD-free (OR [95% CI]: 1.45 [1.38 – 1.52]; p = 3.9x10^-56^; Supplementary Figure 5) stratified cohorts. rs429358 genetic signals in dementia-adjusted and dementia/AD-free GWAS were replicated in AoU (Supplementary Table 2). The *APOE* genetic signal remained highly significant in an age-stratified GWAS, on participants with age 60 years or more (OR [95% CI]: 1.7 [1.63 – 1.77]; p = 2.7x10^-141^; Supplementary Figure 6 and Supplementary Table 2).

### Multi-trait analysis between delirium and Alzheimer disease

We found a significant genetic correlation (*r_g_*) between delirium and Alzheimer disease (AD), *r_g_* = 0.38 (*SE*: 0.12; *p* = 1.9×10^−3^), pointing out to a partially shared genetic architecture. Given this, we further conducted a multi-trait analysis of GWAS summary statistics (MTAG) between delirium and AD. MTAG can increase statistical power to detect new genetic associations, by leveraging the shared genetic information between related traits^34^. Our MTAG analysis identified 10 independent genetic loci associated with delirium, of which 5 replicated in the held-out set (Figure 3 and Supplementary Table 3). The closest genes mapped to the lead replicated variants included: *CR1* (rs4844610 A>C; OR [95% CI]: 1.01 [1.006 - 1.014]; p = 1.4x10^-8^), *BIN1* (rs6733839 T>C; OR [95% CI]: 1.015 [1.012 - 1.018]; p = 7x10^-25^), *CLU* (rs2279590 T>C; OR [95% CI]: 0.992 [0.989 – 0.995]; 4.2x10^-8^), *MS4A4A* (rs1582763 A>G; OR [95% CI]: 0.991 [0.988 - 0.994]) and *TOMM40* (rs117310449 T>C; OR [95% CI]: 1.09 [1.07 - 1.1];p =7x10^-38^).

**Figure 3:**
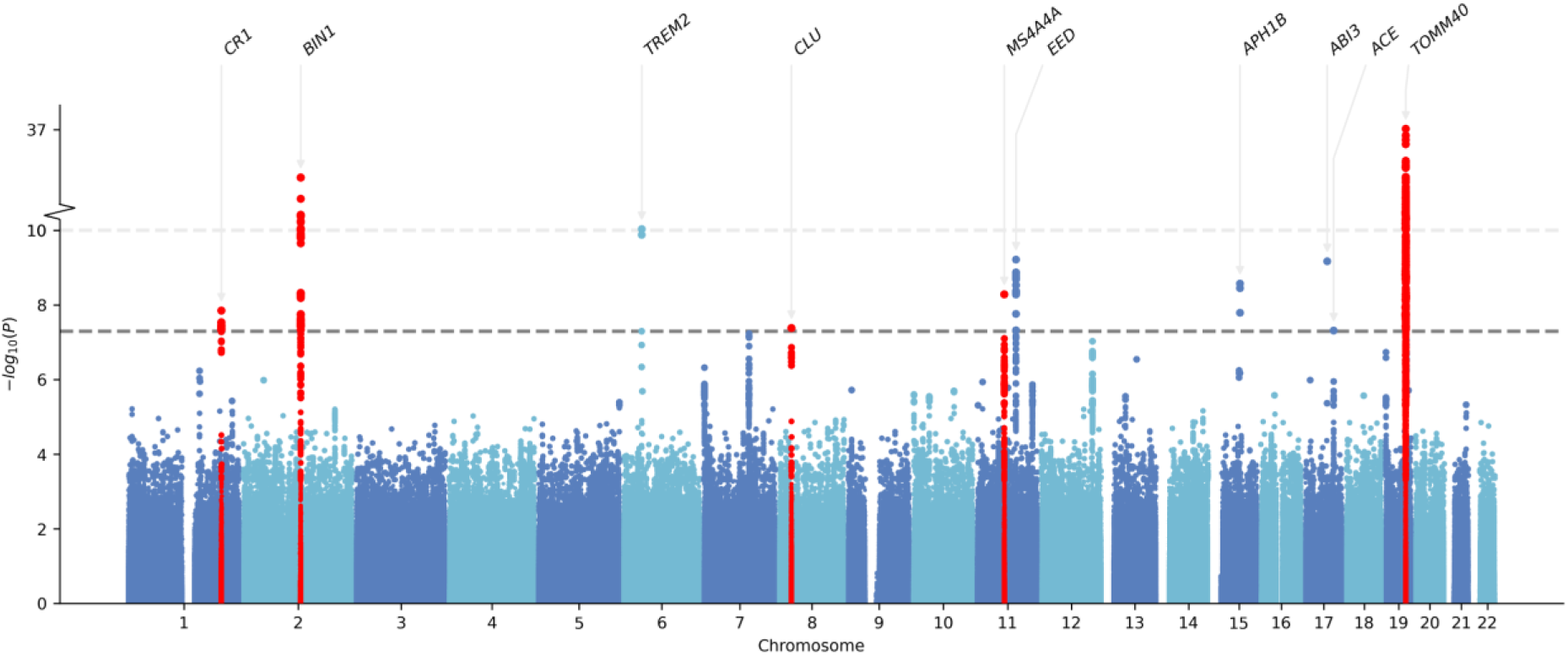
Manhattan plot of the multi trait analysis of GWAS between delirium and Alzheimer disease. Delirium-specific MTAG summary statistics are shown. Each point represents a genetic variant. The x-axis denotes the variant’s genomic position and the y-axis the MTAG association p-value (two-sided) in the discovery set. The grey dashed line denotes the genome-wide significance p-value threshold 5x10^-8^. The closest gene in which the lead significant variant is located is annotated. Red colour highlights loci ±1,000kb around lead variants that replicated. MTAG: Multi-trait analysis of GWAS. The plot was created using the GWASLab python package.

Four out of five of the lead replicated MTAG signals were also significant in the original delirium GWAMA (Supplementary Table 3), with rs2279590 (*CLU* gene) being marginally non-significant (p = 0.058). Effect sizes were consistent between MTAG and delirium GWAMA results. Variants in the *APOE* genomic region that were significant in our delirium GWAMA were also significant in MTAG.

### Mediation analysis

We implemented a mediation analysis in order to estimate how much of the *APOE-*ε4 genetic effect on delirium is mediated by dementia, adjusted for age and sex (Figure 4). We found that *APOE-*ε4 exerts a significant direct effect on delirium in a dose-dependent way: OR _direct effect (0vs1)_ [95% CI]: 1.14 [1.08 - 1.2]; p = 4.5x10^-7^; for one *APOE-*ε4 copy, and OR _direct effect (0vs2)_ [95% CI]: 1.29 [1.15 - 1.45]; p = 1.7x10^-5^ for two *APOE-*ε4 copies. The *APOE-*ε4 total effect on delirium was partially mediated by dementia (OR _indirect effect (0vs1)_ [95% CI]: 1.39 [1.36 - 1.42]; p = 1.1x10^-237^; and OR _indirect effect (0vs2)_ [95% CI]: 2.59 [2.47 – 2.77]; p = 2.1x10^-233^), with the direct effect accounting for 29% and 21% of the total effect for one and two *APOE-*ε4 copies respectively. These results further support the role of *APOE* gene on delirium, independently of dementia.

**Figure 4:**
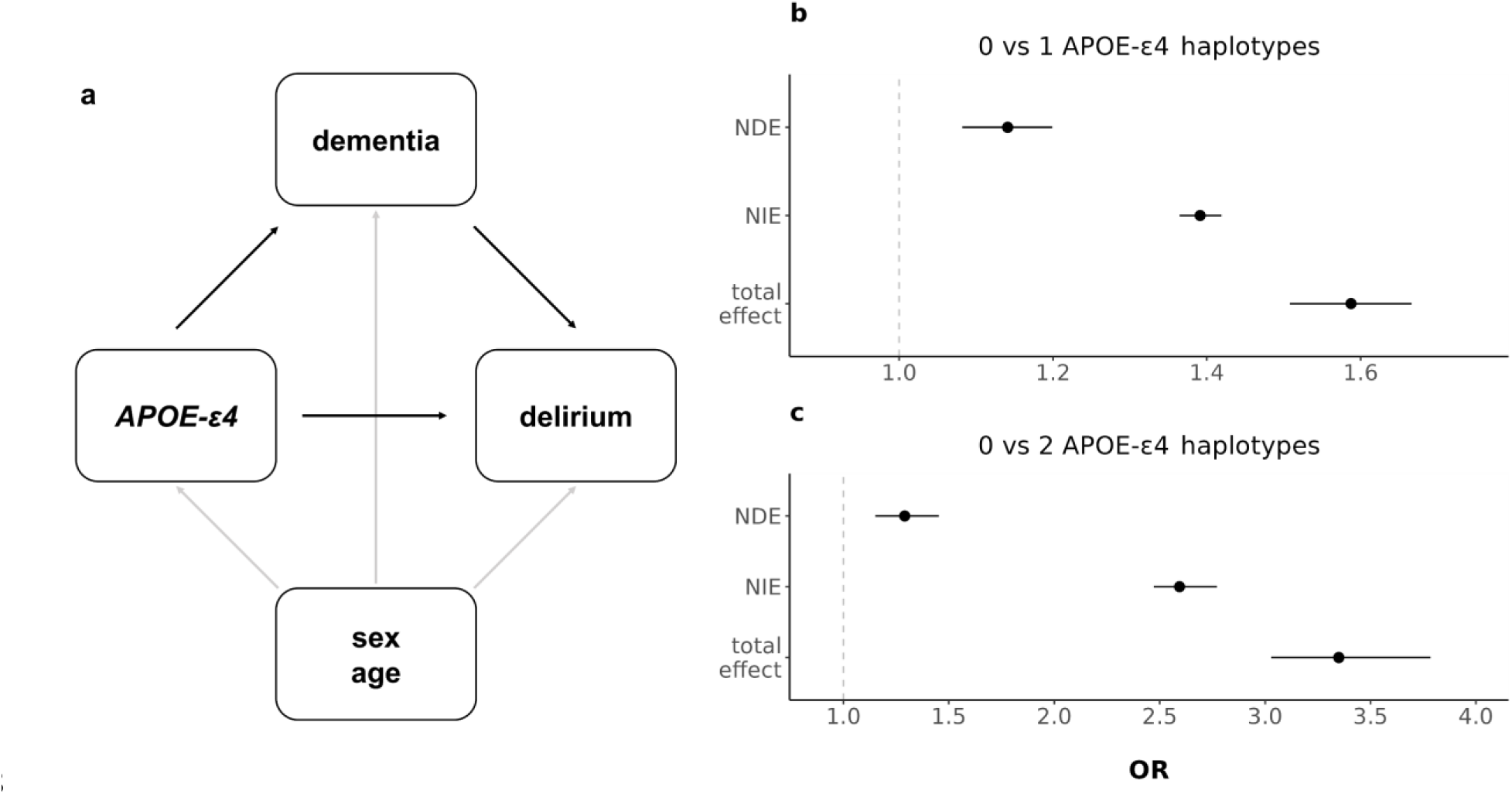
Mediation analysis of APOE-ε4 on delirium, mediated by all-cause dementia. **(a)** Hypothesised directed acyclic graph, showing all-cause dementia as mediator of APOE-ε4 genetic effect on delirium (black arrows), adjusted for baseline covariates: age and sex (grey arrows). **(b,c)** Effect decomposition of the total APOE-ε4 effect, into natural direct (independent of dementia) and natural indirect (mediated through dementia) effects. **(b)** shows the effect of having one APOE-ε4 haplotype, and **(c)** the effect of having two APOE-ε4 haplotypes, compared to having none. OR: odds ratio. NDE: natural direct effect; NIE: natural indirect effect.

Our sensitivity analysis regarding the role of unmeasured confounding suggests that, an unmeasured confounder associated with both dementia and delirium with approximate effect sizes of 1.54 each, over and above the measured covariates, could suffice to completely explain away the observed direct effect of one *APOE-*ε4 copy on delirium, but weaker confounding could not^36^. Respectively, the minimum unmeasured confounder effect size would need to be 1.9 to completely explain away the direct effect of two *APOE-*ε4 copies.

Finally, a mediation analysis further adjusting for cognitive function, socioeconomic deprivation and chronic disease burden, on top of age and sex, still detected a significant direct effect of *APOE-*ε4 on delirium: OR _direct effect (0vs1)_ [95% CI]: 1.12 [1.01 - 1.25]; p = 0.038 (32% of the total effect) for one *APOE-*ε4 copy, and OR _direct effect (0vs2)_ [95% CI]: 1.55 [1.18 - 2.06]; p = 2x10^-3^ (34% of the total effect) for two *APOE-*ε4 copies respectively (Supplementary Figure 7).

### Protein risk factors for incident delirium

A proteome-wide association study (PWAS)on 32,652 European UKB participants (541 cases) revealed 109 out of the 2,919 total proteins, whose plasma levels were significantly associated with incident delirium up to 16-years of follow-up (Figure 5a and Supplementary Table 4) at a Bonferroni adjusted p-value threshold (p = 1.7x10^-5^). The APOE protein had a negative effect on incident delirium risk, meaning that higher plasma levels of the protein are associated with reduced future risk. The association was significant at the nominal level (0.05), but not after multiple test correction (OR [95% CI]: 0.86 [0.79 - 0.94], p = 7x10^-4^, Supplementary Table 4).

**Figure 5:**
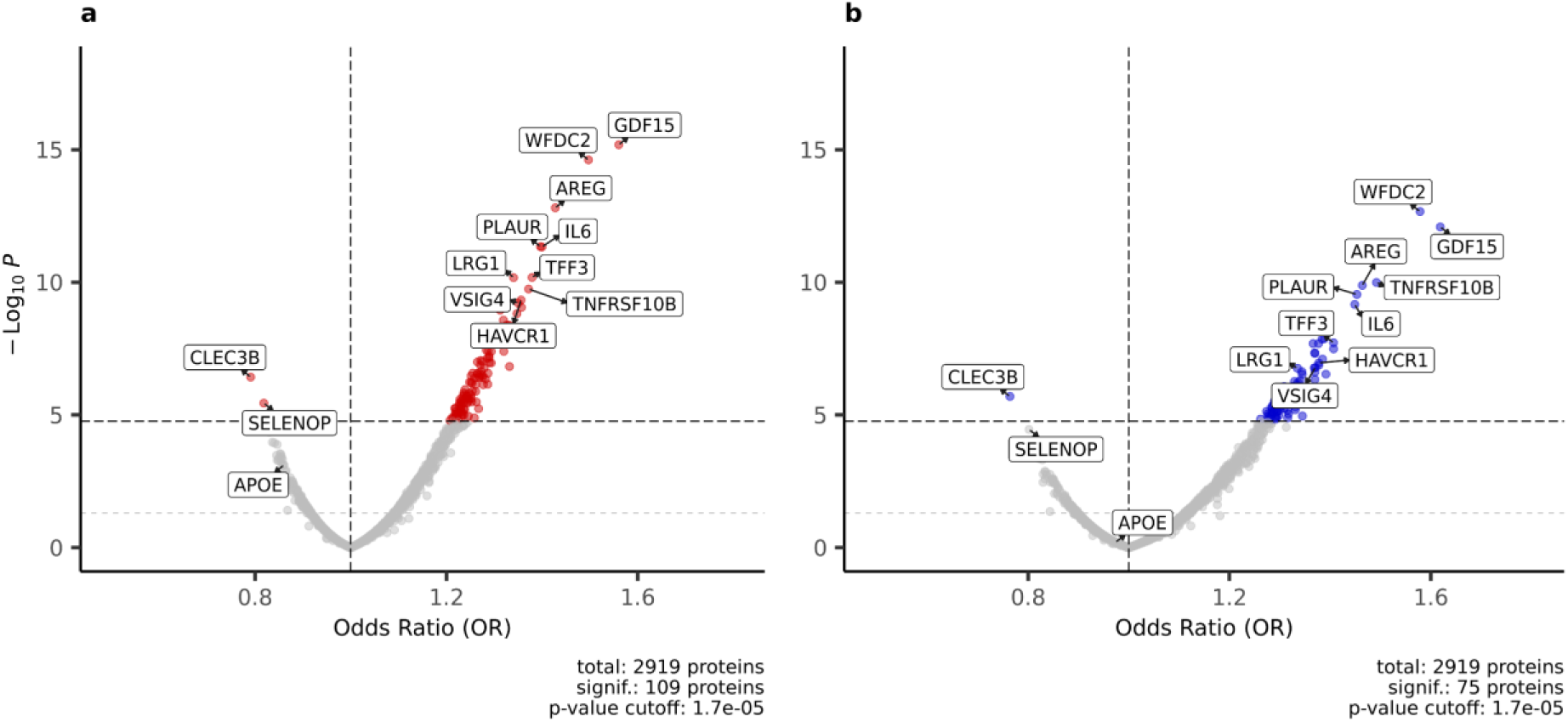
Associations of plasma proteins with incident delirium. Volcano plot showing the proteome-wide associations of 2,919 plasma proteins with incident delirium in (**a**) the non-stratified, and (**b**) the dementia-free UKB proteomic set. Odds ratios of the logistic regression models are plotted on the x-axis and respective two-sided -log_10_(p-values) on the y-axis. All models are adjusted for age at protein collection, sex and BMI. The black dashed horizontal line indicates the Bonferroni adjusted p-value threshold: 1.7x10^-5^, with proteins above this line colored red. The grey dashed horizontal line indicates the nominal p-value threshold: 0.05. The plot was created using the EnhancedVolcano R package (v1.18.0).

Further adjusting the individual protein models for *APOE-*ε4 status did not substantially alter the results (Supplementary Table 5), having highly correlated effect size estimates with the models adjusted only for age, sex and BMI (Pearson’s correlation r=0.99). Additionally including a protein* *APOE-*ε4 interaction term also yielded highly similar results (main effect correlation r=0.93). No significant protein*\* APOE-*ε4 interaction has observed at a Bonferroni adjusted threshold of p-value < 1.7x10^-5^ (Supplementary Table 6).

The 109 proteome-wide significant proteins were found to be significantly enriched (q-value < 0.05) in several important inflammation and immune response biological pathways, such as interleukin and Tumour Necrosis Factor (TNF) signalling (Supplementary Table 7).

Restricting our PWAS on the dementia-free subset of UKB (31,692 participants; 349 delirium cases), reduced the number of proteins significantly associated with incident delirium to 75 (Figure 5b and Supplementary Table 8). Notably, the APOE plasma protein, although still had a negative effect size, lost nominal significance and had reduced magnitude that in the non-stratified PWAS (OR [95% CI]: 0.97 [0.87 – 1.08]; p = 0.63).

### Protein selection and prediction models

A machine learning framework was applied to further pin down which plasma proteins are robustly associated with incident delirium. Our approach, based on LASSO regularised regression^37^ and stability selection^38^, revealed 19 proteins (stability-selected proteins, Supplementary Table 9) consistently selected as predictive of incident delirium in the training set. All of the stability-selected proteins represent a subset of the top individually significant proteins identified through the proteome-wide association analysis. The FGL1 protein was removed from subsequent analyses, as it had a non-significant contribution to the re-fit prediction models and was dropped during stepwise regression. The 18 remaining stability-selected proteins provide marginal prediction improvements of incident delirium on an independent test set, compared to predictions based on demographic factors alone (Figure 5 and Supplementary Tables 10-11). Specifically, adding the 18 stability-selected proteins to the “basic” model that includes age, sex and BMI as predictors increased the AUC from 0.764 to 0.791, but the increase was not significant (DeLong test p-value = 0.09, Supplementary Table 11). However, adding proteins and *APOE-*ε4 status to the basic model showed a significant prediction improvement (AUC from 0.764 to 0.794, p = 0.049, Supplementary Table 11). Finally, the model fit only with the selected proteins performed worse that the basic model (AUC from 0.764 to 0.729, p = 0.21, Figure 5a and Supplementary Table 11). The precision-recall performance showed similar pattern (Figure 5b), with the full models having higher PR-AUC: 0.065 and 0.06 for the “*APOE*+proteomic+basic” and the “proteomic+basic” models respectively, compared to 0.043 for the basic model.

We repeated our protein selection and prediction framework in the dementia-free sub-cohort of the UKB proteomic set. 14 out of the 19 stability-selected proteins still had a significant effect on incident delirium in the training set (Supplementary Table 9). However, only the AREG and MSLN proteins were consistently selected in the dementia-free sub-cohort (selection frequency > 0.5). We still observed a marginally better prediction of incident delirium on the test set using *APOE-*ε4 and the 14 stability-selected proteins on top of demographic factors (AUC from 0.758 to 0.79, p = 0.13; PR-AUC from 0.033 to 0.036) (Supplementary Figure 8 and Supplementary Table 10), although now with a non-significant AUC improvement.

**Figure 6:**
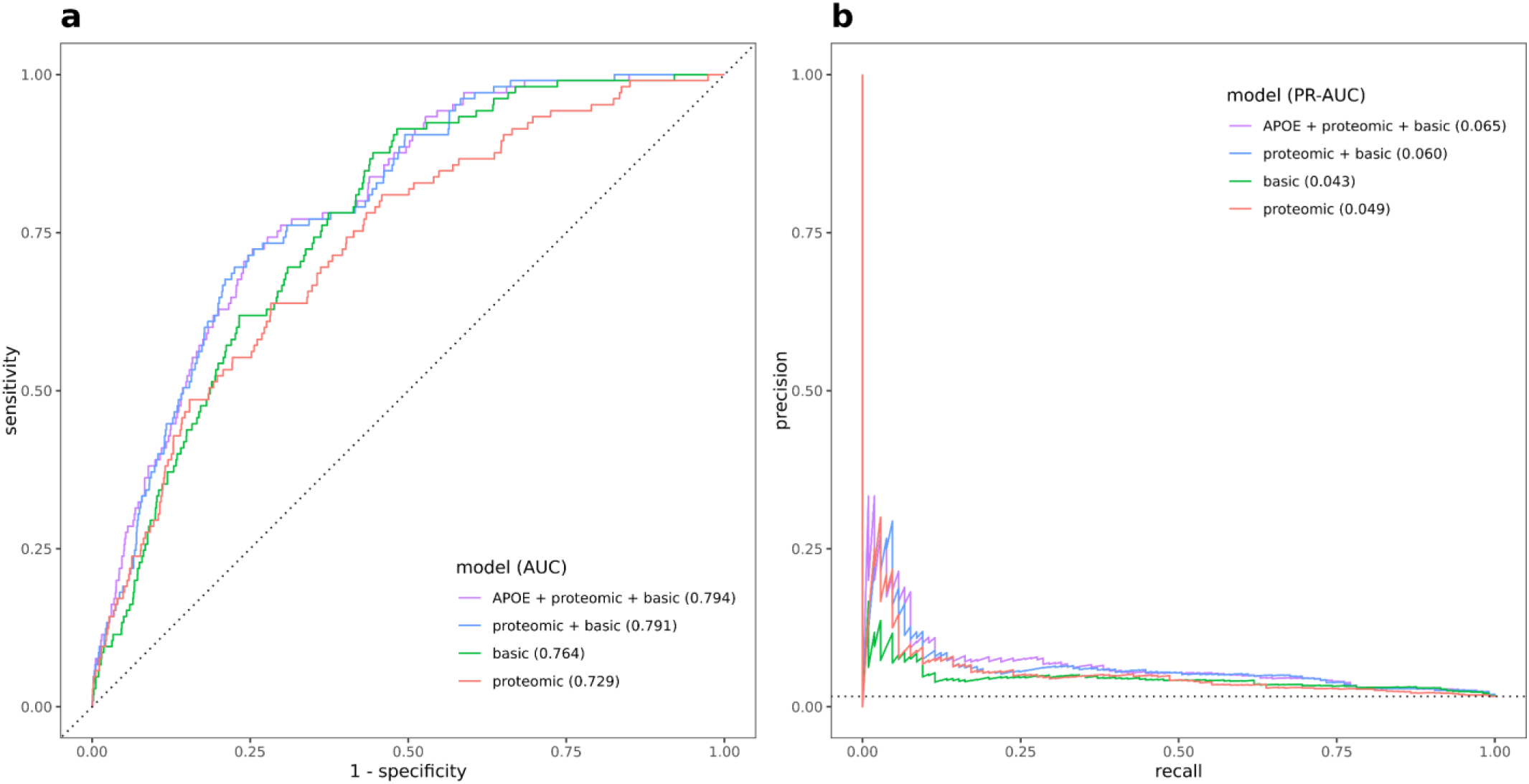
Proteomic predictive performance on test set. Receiver Operating Characteristic (ROC) (**a**) and Precision-Recall (PR) curves (**b**) showing performance metrics of different models for predicting incident delirium on the held-out test set. The “proteomic” logistic regression model is fit with the 18 stability-selected proteins, the “basic” model is fit using only age at protein collection, sex and BMI and the “proteomic+basic” includes both. The “APOE+proteomic+basic” model additionally includes APOE-ε4 haplotype count as predictor. The dotted black lines indicate the performance of a randomly classifying model, that is the diagonal of the ROC curve (**a**) and the disease prevalence horizontal line in the PR curve (**b**). AUC: Area Under the Curve. ROC and PR plots and AUC calculations are made using the yardstick R package (v1.3.0).

### Mendelian randomisation

We performed mendelian randomisation (MR), to test for causal associations between 1,989 plasma proteins (exposures) and delirium (outcome). We found 53 proteins causally associated with delirium at a false discovery rate (FDR) q threshold < 0.05 using inverse variance weighted (IVW) or Wald ratio MR. (Figure 7 and Supplementary Table 12). Out of the 53 MR significant proteins, DSC2 was the only one with a significant association in our PWAS analysis of protein risk factors (Figure 5) and a consistent direction of effect. Using a less strict FDR q < 0.05 threshold in our PWAS, 7 additional proteins were identified (ADAM8, APOE, DPP10, DSC2, LAYN, NT5C1A, PILRA and PVR) that were significant in both the PWAS and MR. At a nominal PWAS p-value level (p < 0.05), three more proteins (CCL25, PON3 and GGH) overlapped the significant MR results. Four of the above PWAS - MR overlapping significant proteins successfully replicated in our FinnGen replication MR: GGH, PVR, APOE and PILRA (Figure 7). All of the replicated delirium-associated proteins were also significant using sensitivity MR methods (weighted median, maximum likelihood and MR-Egger; p < 0.05) (Supplementary Table 13). The MR-Egger intercept test did not reveal evidence of horizontal pleiotropy (p > 0.05). Genetic variant heterogeneity was suggested by the Cochran’s Q test (p < 0.05) for the genetic instruments used in the APOE and PVR MR (Supplementary Table 13). Repeating MR for these two proteins using more strictly independent pQTLs (r^2^ < 0.001), APOE did not show evidence for heterogeneity (Q test p = 0.37), while its causal effect on delirium remained significant (IVW MR beta (standard error) = -0.57 (0.03), p = 1.13x10^-78^). For PVR, heterogeneity remained high (Q test p = 0.03), and lost significance for its causal effect (IVW MR beta (SE) = 0.06 (0.06), p = 0.34), suggesting that PVR’s initially observed significant MR effect may be driven by pleiotropy and linkage disequilibrium of its genetic instruments with the *APOE-*ε4 haplotype that is located in close proximity. Using delirium in the dementia-free UKB GWAS as outcome, ADAM8, CCL25, PILRA, and LAYN lost their MR causal effect significance (FDR q > 0.05). A summary of analyses supporting each protein’s association with delirium is presented in Table 1, with full summary results in Supplementary Data 1.

**Figure 7:**
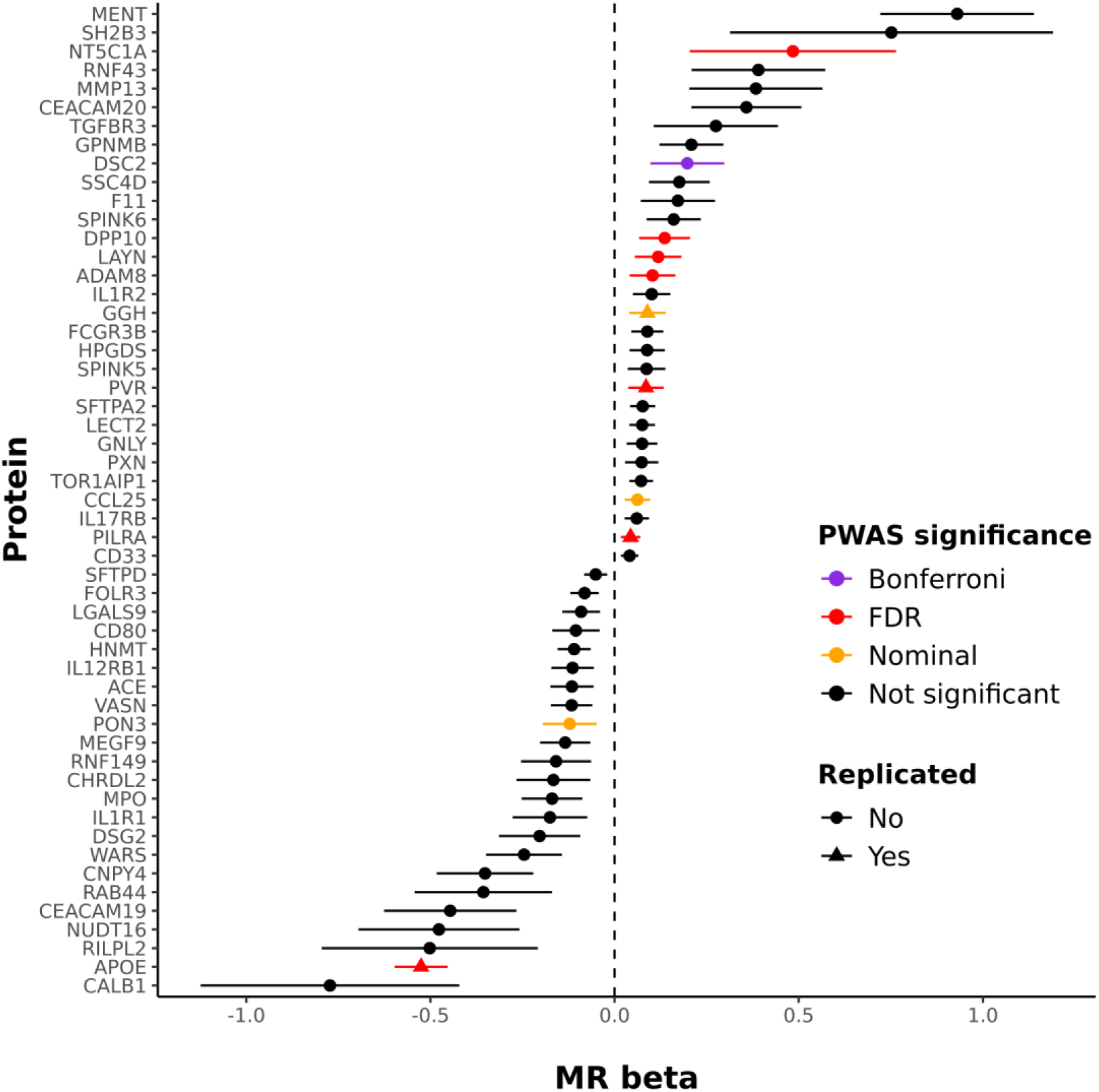
Mendelian randomisation results of plasma proteins on delirium. MR associations were derived using the IVW or Wald ratio method and UKB delirium GWAS summary statistics. Only significant (FDR q < 0.05) MR associations are shown (IVW/Wald ratio betas and 95% confidence intervals). Replication MR was conducted using delirium GWAS summary statistics from Finngen. Colors are assigned to proteins based on their significance in the PWAS analysis: Bonferroni: p < 1.7x10^-5^; FDR: q < 0.05; nominal: p < 0.05 and having consistent direction of effect. MR: mendelian randomisation; PWAS: proteome-wide association study; FDR: false discovery rate. IVW: inverse variance weighted.

**Table 1:** Overview of follow-up analyses on proteins significantly associated with delirium through PWAS and MR.

| Protein | Effect direction | PWAS significance | Discovery MR (UKB) |  |  |  |  |  |  | Replication MR IVW (FinnGen) | Co-localisation | Druggability Tier |
| --- | --- | --- | --- | --- | --- | --- | --- | --- | --- | --- | --- | --- |
|  |  |  | IVW | IVW dementia-free | Weighted median | Maximum Likelihood | MR-Egger | MR-Egger intercept test | Cochran's Q test |  |  |  |
| PON3 | - | Nominal | ✓ | ✓ | ✓ | ✓ | ✓ | ✓ | ✓ | ✗ | ✗ | Tier 1 |
| ADAM8 | + | FDR | ✓ | ✗ | ✓ | ✓ | ✗ | ✓ | ✓ | ✗ | ✗ | Tier 2 |
| GGH | + | Nominal | ✓ | ✓ | ✓ | ✓ | ✓ | ✓ | ✓ | ✓ | ✓ | Tier 2 |
| PVR <sup>a</sup> | + | FDR | ✓ | ✓ | ✓ | ✓ | ✓ | ✓ | ✗ | ✓ | ✗ | Tier 3A |
| APOE <sup>b</sup> | - | FDR | ✓ | ✓ | ✓ | ✓ | ✓ | ✓ | ✗ | ✓ | ✓ | Tier 3A |
| DPP10 | + | FDR | ✓ | ✓ | ✓ | ✓ | ✗ | ✓ | ✓ | ✗ | ✗ | Tier 3B |
| CCL25 | + | Nominal | ✓ | ✗ | ✓ | ✓ | ✓ | ✓ | ✓ | ✗ | ✗ | Tier 3B |
| DSC2 | + | Bonferroni | ✓ | ✓ | ✓ | ✓ | ✗ | ✓ | ✓ | ✗ | ✗ | Not classified |
| NT5C1A | + | FDR | ✓ | ✓ | ✓ | ✓ | ✗ | ✓ | ✓ | ✗ | ✗ | Not classified |
| PILRA | + | FDR | ✓ | ✗ | ✓ | ✓ | ✓ | ✓ | ✓ | ✓ | ✗ | Not classified |
| LAYN | + | FDR | ✓ | ✗ | ✗ | ✓ | ✗ | ✓ | ✓ | ✗ | ✗ | Not classified |

Proteins that were supported by both the PWAS ( $p < 0.05$ ) and MR via IVW or Wald ratio ( $FDR < 0.05$ ) analyses in UKB and had consistent effect direction are shown. ✓ (green) denotes significance ( $p < 0.05$ ) in the MR sensitivity analyses (Weighted median, Maximum Likelihood and MR-Egger); non-significance in the MR-Egger intercept and Cochran's Q tests ( $p > 0.05$ ); and $PP.H4 > 0.8$ in the colocalisation analysis. ✓ (orange) denotes suggestive colocalisation: $0.8 >$ $PP.H4 > 0.5$ ✗ denotes failure. Full summary statistics are available in Supplementary Data 1. PWAS: proteome-wide association study; MR: mendelian randomisation; FDR: false discovery rate; IVW: inverse variance weighted; UKB: UK Biobank; pQTL: protein Quantitative Trait Loci
<sup>a</sup> PVR lost MR significance and showed high heterogeneity using strictly independent pQTL ( $r^2 < 0.001$ ).
<sup>b</sup> APOE remained significant in MR and did not show heterogeneity evidence using strictly independent pQTL ( $r^2 < 0.001$ ).

### Colocalisation

We further conducted colocalisation analysis to test whether the protein and delirium genetic effects are exerted by the same rather than distinct genetic variants in each genomic region. 37 proteins colocalised with delirium within the *APOE* genomic region (Supplementary Table 14 and Supplementary Data 2) with high probability (posterior probability of a shared causal variant, PP.H4 > 0.9 as per coloc software notation^39^). 30 more proteins showed suggestive evidence of colocalisation with delirium throughout the genome (PP.H4 > 0.5). It should be noted that colocalisation methods are generally not well powered to detect shared casual genetic signals unless both traits shown strong genetic support (i.e. GWAS p < 5x10^-8^)^39,40^. Here, only the *APOE* region is strongly associated with delirium, thus many colocalisation signals in other parts of the genome may be missed.

### Druggability assessment

For delirium-associated proteins with support from both our PWAS and MR analyses, we assessed their suitability to act as drug targets (Table 1). We used the “druggable genome” curated by Finan et al (2017)^41^, a resource which stratified human proteins on tiers based on the strength of their druggability evidence (see Methods). Of the selected proteins, PON3 belonged in the druggability tier 1 (targets of approved drugs or clinical-phase drug candidates); ADAM8 and GGH belonged in tier 2 (“targets with known bioactive drug-like small-molecule binding partners”^41^ or having high similarity with approved drug targets); PVR, APOE, DPP10, CCL25 in tier 3 (secreted or extracellular proteins, proteins with medium similarity to approved drug targets, and members of key druggable gene families not included in tiers 1 or 2^41^). DSC2, NT5C1A, PILRA, LAYN did not have druggability information.

### KGWAS

Knowledge Graph GWAS (KGWAS) analysis of our GWAMA summary statistics revealed 4 disease-critical genes within the *APOE* genomic region (*APOE*, *TOMM40*, *PVRL2*, *BCAM*), prioritized via functional genomics knowledge graph^42^. All of the KGWAS-identified significant SNPs were also significant in our GWAMA (Supplementary Table 15).

## Discussion

In this analysis we conducted the largest to our knowledge multi-ancestry genome-wide meta-analysis on delirium. Genetic variants on the *APOE* gene on chromosome 19 were identified as significantly associated with delirium, with the top variant hit, rs429358, showing population-specific association patterns. The *APOE* gene encodes for APOE, a lipid transporter protein in the periphery and the brain. APOE is a strong risk factor for Alzheimer Disease (AD), through its diverse roles in pathways such as amyloid-β plaque deposition, neuroinflammation and dysregulation of lipid metabolism in the brain^43^.

The role of *APOE* gene in delirium is currently unclear, with previous meta-analyses reporting no association between *APOE* and delirium^12,23^. In UKB, a previous study on European participants found an association between *APOE-*ε4 status and delirium (hazard ratio = 3.73 [2.68 - 5.21])^44^. It has been suggested that interactions between *APOE-*ε4 and inflammation-related proteins can drive delirium development^6^. To assess this hypothesis, we tested whether the interaction term between each plasma protein level and *APOE-*ε4 status significantly associated with incident delirium (Supplementary Table 5). No protein\**APOE-*ε4 interaction reached significance adjusted for multiple testing (p-value threshold = 1.7x10^-5^). However, the CEND1 protein exhibited an interaction with *APOE-*ε4 marginally below threshold (beta_(interaction)_ [95% CI] = 0.27 [0.10 – 0.42]; p_(interaction)_ = 8x10^-4^). CEND1 is a mitochondrial neural differentiation protein, expressed in the nervous system^45^. It has previously been implicated in cognitive impairment in mice^45^ and AD in human^46^ brains. Additionally, *APOE-*ε4 expression in astrocytes has been implicated in impaired mitochondrial function^47^, although to our knowledge CEND1 and APOE have not been linked in previous studies. CEND1 role in delirium has also not been investigated so far.

Ancestry-dependent *APOE-*ε4 genetic effects on delirium have not been systematically assessed previously. For AD, the risk conferred by *APOE-*ε4 varies by ancestral background, with African/African Americans and Hispanics having less pronounced risk than white Europeans and Asians^48,49^. Additionally, higher *APOE-*ε4 expression levels have been observed in carries of European compared to African ancestry^50^. Here, findings suggest a similar pattern, that is rs429358-C having a higher effect in European, Finnish and south Asian populations than African and Hispanic/Admixed American. Overall, sub-populations from the USA (AoU, MGI) have weaker *APOE* effects than those from the UK or Finland (UKB, FinnGen). This might reflect the younger age of participants in USA-based studies (Supplementary Table 1) or phenotypic differences in delirium diagnoses across the healthcare systems of different countries. Age-dependent genetic effects have been previously described for *APOE-*ε4 with regard to AD^48,51^ and progression to mild cognitive impairment and AD^52^, showing increasing effects until an age of 70 – 75, with reduced effect on later ages^48,51,52^.

It is also possible that underlying dementia is driving the strong association between *APOE* and delirium observed here. 36% of UKB European delirium cases had a dementia diagnosis, compared to 1.4% in the control group (Supplementary Table 1). This is to be expected given the close relationship of the two disorders^6^, but it may hide delirium specific genetic effects and overemphasize the role of *APOE*. To this end, *APOE* remained significant in our range of sensitivity analyses – GWAS adjusting for all-cause dementia (Supplementary Figure 3), mediation analysis (Figure 4), dementia and AD stratified GWAS (Supplementary Figure 4-5 and Supplementary Table 2) – although with weaker effect. This result may suggest that *APOE* association with delirium is not entirely through its role in dementia or AD.

Regarding our mediation analysis, it should be noted that unmeasured confounding could bias the estimate of *APOE’s* direct effect on delirium^36,53^ (see Methods section). Unmeasured confounding between *APOE* (exposure) and dementia (mediator) or delirium (outcome) could be introduced by residual population stratification or linkage disequilibrium between *APOE-*ε4 and linked causal genetic variants. It is also possible that *APOE*-induced pleiotropic genetic effects can confound the delirium-dementia relationship, also introducing bias. Our results remained significant after adjusting for possible confounders between dementia and delirium, i.e. shared risk factors such as cognitive function, socioeconomic deprivation and comorbidity burden^3–5,54,55^. None of the additional confounders had an independent odds ratio effect on both delirium and dementia above 1.54 each, the minimum required to explain away the observed *APOE* direct effect on delirium. However, some residual confounding may still remain, for example due to precipitating factors of delirium episodes during hospitalisation, such as infections, medication status, sleep disruptions or severity of presenting illness. These factors have been previously estimated to have a strong effect on delirium^3,4^, and may be also related to dementia^55^. Assessing the effect of those factors was outside the scope of our study.

Adjusting for *APOE-*ε4 attenuated the genetic effects within the whole *APOE* region. This observation suggests that the significance of the genetic variants in the close proximity is driven by linkage disequilibrium with the *APOE-*ε4 haplotype, not secondary independent signals. Moreover, an intronic variant on the *ADAM32* gene gained significance after adjusting for *APOE-*ε4. *ADAM32* belongs to the ADAM family of metalloproteinases, some of which have been implicated in AD^56^. ADAM proteins are involved diverse functions, including immunity related pathways^57^.

Regarding our multi-trait analysis of delirium with AD (MTAG), five genetic loci were found to have a significant effect on delirium, supported by replication in the AoU EUR cohort. Among the replicated loci, several important AD risk genes^58^ were detected: *BIN1*, *CLU*, *CR1*, *MS4A4A*, *TOMM40*. The *MS4A4A* gene is expressed in macrophages and has been linked with AD^59^, vascular dementia and systemic lupus erythematosus^60^. The lead variant’s minor allele on the *MS4A4A* gene, rs1582763, has been previously associated with decreased risk of AD^59^. This variant’s association with delirium has not been reported before, but we also found a protective effect of the rs1582763 minor allele on delirium. *BIN1* has been recently implicated in the regulation of calcium homeostasis in glutamatergic neurons, and its expression in AD human brains is reduced comparted to healthy brains^61^. The clusterin gene (*CLU*), also named Apolipoprotein J (*APOJ*) codes for a multifactorial protein, with apparent role in neurodegenerative diseases^62^. *CLU*, much like *APOE* is thought to be involved in amyloid-β plaque deposition in AD pathologies. With regard to delirium, protein expression of apolipoproteins including CLU and APOE were previously found to be downregulated in the cerebrospinal fluid (CSF) of delirium subjects compared to mild AD controls^63^. In our proteomic analysis, CLU protein levels were also downregulated in incident delirium subjects’ plasma (Supplementary Table 3), but not significantly so (p-value > 0.05). This discrepancy may reflect different CLU protein abundance between CSF and plasma tissues. The *CR1* gene, implicated in complement activation, is believed to exert its role to AD pathogenesis through amyloid-β clearance, neuroinflammation and tauopathy (the deposition of abnormal tau protein in the brain)^64^. To the best of our knowledge, the role of *CR1* in delirium has not been investigated previously. In our study, CR1 plasma protein levels had a nominally significant association with incident delirium (p-value = 0.013). *TOMM40* genetic variants have been associated with AD before^65^. Given *TOMM40*‘s close proximity and high linkage disequilibrium with the *APOE* gene, its role in AD has frequently been contested^65^. However, it has also been suggested that *TOMM40* independently affects AD risk through its role in regulating protein transportation in the mitochondria^65,66^. *TOMM40* has not been previously implicated in delirium. Overall, given the small effect sizes of the MTAG-identified genes for delirium (Supplementary Table 2), and their prominent role in AD, it may be possible that their significance in our analysis is mainly driven due to their role in AD. Nonetheless, our findings suggest novel genes (e.g. *MS4A4A*, CR1, *TOMM40*) that may be of relevance to future delirium research and therapeutic targets investigations.

Previous proteomic studies for delirium have, to our knowledge, focused on the protein landscape shortly before or following delirium episodes^31^. Here, an important novelty of our proteomic study is the large follow-up period between protein measurements and delirium incidence (median time to first delirium event: 11.4 years, interquartile range (IQR) [9.5-12.7]), allowing identification of relevant biomarkers and biological mechanisms at an early stage, such as several neurologically relevant and immune system related proteins.

For example, we found plasma GFAP and NEFL levels – biomarkers of neuronal injury – to be associated with incident delirium. Previous research paints a complex, possibly bidirectional relationship between neuronal injury and delirium. Increased plasma and CSF NEFL levels, a marker of neuroaxonal injury, have been observed in delirium patients pre- and postoperatively^6,8,9,67,68^. It is unclear however whether this is due to underlying neurodegeneration (e.g. due to preclinical dementia) or a feature of delirium^6^. In our proteomic analyses we observed significant and consistent effect of NEFL on incident delirium in both the full and dementia-free populations. This may implicate neuroaxonal injury as an underlying driver of delirium independently of dementia-related neurodegeneration, even long before hospitalisation. Our findings, thus, add support for the role NEFL as a useful predictive biomarker for delirium^6^.

GFAP is an astrogliosis marker, that is an increase of astrocytes levels in the central nervous system in response to injury. High GFAP levels have been observed previously in post-mortem brains of delirium cases^69^, CSF of persistent delirium^8^, blood and CSF of delirium patients undergoing elective surgery^32,67^. In our analysis, GFAP, was only predictive of incident delirium in our full proteomic set (Supplementary Table 4), not in the dementia-free population (Supplementary Tables 8-9). This may indicate that, in the years preceding delirium, astrogliosis is only relevant for delirium in the context of underlying dementia. GFAP may be more useful as a delirium biomarker in the short term, possibly in patients with traumatic brain injury or underlying neurodegeneration.

Systemic inflammation markers have been observed among the delirium-associated proteins in our study. For instance, C7, BTLA, FGL1 participate in the immune response, whereas LRG1 and LTA4H in inflammatory processes. LRG1 has been implicated in brain injury after sepsis in mice^70^, sepsis being a main driver of delirium aetiology^3^. Interleukins (IL), identified through our enrichment analysis, play an important role in regulating immune response and inflammation and have frequently been implicated in delirium^71,72^. Indeed, IL-6 – found to have a strong delirium association in our proteome wide association study – is among the most consistently identified biomarkers of delirium and markers of postoperative cognitive decline^31,32^. All of the inflammatory related proteins were significant in our dementia-free sensitivity analyses, suggesting a role for delirium partially independent of dementia pathology.

Other notable proteins robustly associated with delirium in our proteomic analysis are BCAN, SELENOP, AREG and MSLN. BCAN, a protein with a role in brain extracellular matrix formation, has been observed to be downregulated in brains of post-infection delirium and in AD patiens^73^. Lower plasma levels of SELENOP, an important selenium transporter in the brain has been associated with worse global cognition and AD^74^. Here, to the best of our knowledge, is the first time that an association between SELENOP and delirium has been reported. The associations of AREG and MSLN with delirium were also novel findings. AREG is a member of epidermal growth factor protein family, involved in many aspects of cell proliferation^75^. Possible mechanisms linking AREG to delirium could include neuroinflammation and regulation of T cells in astrocytes in sepsis-associated delirium^76^, type 2 inflammatory response and tolerance against pathogens^77^, or tumor development^75^. MSLN is a mesothelial cell surface glycoprotein, overexpressed in many cancers^78^. Delirium is common in cancer patients, arising either as a complication of the disease or indirectly through medical and surgical intervention for cancer treatment^79^. Further research on the overlapping pathophysiology between delirium and cancer (e.g. shared biomarkers^80^) is important for better management of delirium throughout cancer care trajectories^79^.

Overall, our proteomic analysis results align with the proposed mechanisms of delirium pathophysiology, that is brain vulnerability - indicated by brain injury marker proteins -, systemic and nervous system inflammation being driving factors for delirium^3,6^. At the same time, our results could inform future research in delirium prediction biomarkers, some of which are novel in delirium research (e.g SELENOP, CEND1, AREG and MSLN).

Our findings have notable clinical and biological clinical implications. We found that *APOE-*ε4 is a robust genetic risk factor, remaining significant after adjustment for presence of dementia. The findings suggest that *APOE-*ε4 confers vulnerability to adverse acute brain changes induced by triggers of delirium. Indeed, it is known that *APOE-*ε4 is associated with exaggerated neuroinflammatory responses^81^, altered blood brain barrier integrity^82,83^, increased beta-amyloid accumulation following various injuries^84^, and increased white matter pathology^85^. *APOE-*ε4 therefore could contribute to increased risk of delirium, and also cause or amplify neuropathological changes linked with dementia. A key next step is to test whether *APOE-*ε4 carriers are disproportionately likely to develop new dementia after a delirium episode, even in the absence of prior cognitive impairment.

Clinically, *APOE-*ε4 status has potential to assist in stratification for delirium prevention trials, in prediction of dementia following delirium, and as a target for neuroprotective therapies during acute illness especially if delirium is present. Animal studies targeting APOE have shown encouraging results in terms of improving AD phenotypes^83^. Some promising therapeutic approaches include increasing APOE levels and lipidation, APOE mimetics (small peptides mimicking APOE’s receptor-binding structure) and *APOE-*ε4 gene therapy^43,83^. However, therapeutic translation in human clinical trials is still limited^83^. A currently unpublished phase 2 clinical trial (NCT03802396)^86^ found that administration of CN-105 – an APOE mimetic drug – lowered postoperative delirium incidence and severity compared to administration of placebo, but not significantly. Further clinical research is needed in order to clarify the therapeutic potential of APOE targeting therapies.

In terms of delirium treatment, we explored whether our identified proteins could be suitable drug targets. For this, we triangulated our proteomic study findings with proteome-wide mendelian randomisation (MR) and colocalisation analyses. Incorporating genetics in drug target identification, e.g. through MR, is particularity attractive, as drugs with genetic support are more likely to be successful in clinical trials^87,88^ or can offer drug repurposing opportunities^89^. Our druggability assessment found PON3 as an notable drug target for delirium. PON3 is a secreted high density lipoprotein-associated enzyme^90^, participating in the metabolism of the statin lovastatin, a licenced lipid-lowering medication. The role of statins in delirium management is unclear. Statin usage has been associated with improved delirium outcomes in randomised controlled trials and observational studies, but overall evidence is inconsistence, with a recent meta-analysis finding no significant protective effect and high heterogeneity^91^. In our analysis, increased PON3 plasma level was found to have protective effect against delirium. This is consistent with high PON3 levels being protective against atherosclerosis and cardiovascular disease^92^, possibly through its antioxidant action^92,93^. It is not clear, however, how PON3 levels can affect statin metabolism. It may be possible that PON3 can affect lovastatin and other statins’ efficacy via their involvement in the drug activation^90^, which may explain part of the high heterogeneity observed regarding statins’ effect on delirium^91^. Our results may, thus, suggest a reconsideration of a possible role for statins in improving delirium prevention or treatment.

Additionally, we identified proteins previously suggested as therapeutic targets for e.g. cancers (ADAM8^94^, GGH^95^, PILRΑ^96^) and AD (PILRA^97^). This may offer promising opportunities for investigating new delirium treatment targets. However, results should be interpreted with caution, as liberal p-value thresholds – up to p < 0.05 – were used to indicate proteomic support for our druggability assessment, and some proteins (e.g. PON3 and ADAM8) were not replicated in our replication MR, although the same effect direction was still observed (Table 1 and Supplementary Data 1).

The main strength of our analysis is the large-scale investigation of delirium genetic and proteomic risk factors. Both in terms of sample size, follow-up time before protein measurement and delirium episodes, and number of genetic variants / proteins tested, this is to our knowledge the largest study on the molecular background of delirium risk conducted so far. On the other hand, some of the limitations of the study include the underdiagnosis of delirium in hospital health records from which the phenotype was mostly derived^98^. This misclassification could introduce noise to the results, limiting discovery of genetic and protein effects. Also, the small sample sizes of the non-European sub-populations hinders the identification of risk factors specific to them. Moreover, not the full spectrum of the human proteome is captured in the assayed plasma proteins, potentially missing proteins important for delirium biology and prognosis. Finally, although the use of plasma proteins as predictive biomarkers is of great significance, proteomic profiles of delirium-relevant tissues, such as brain or CSF would be invaluable.

In conclusion, our results point out to an oligogenic genetic architecture for delirium, with the *APOE* locus identified as a strong, potentially population specific genetic risk factor, independently of dementia. However, further replication in larger non-European cohorts is required. Our plasma proteome analysis supports previous findings and discovers putatively novel proteins implicated to delirium, some of which suggested for therapeutic applications. Taken together, genetic and proteomic risk factors suggest a shared aetiology between delirium and dementias, possibly contributing to a better understanding of delirium’s complex biological origin and the discovery of clinically relevant biomarkers.

## Methods

### Study populations

The project utilises biomedical data from ancestrally diverse large-scale cohorts. Included cohorts were either (a) databases containing individual-level genomic measurements linked to healthcare records, or (b) previously published summary results from genomic studies on delirium phenotypes. Contributing individual-level cohorts include the UK Biobank^99,100^ (UKB) and the All of Us Research Program^101^ (AoU). Summary results have been obtained from ancestrally Finnish (FinnGen^102^; *n*_*cases*_ = 3,371; *n*_*controls*_ = 388,560) and European participants (Michigan Genomics Initiative cohort (MGI)^103^; *n*_*cases*_ = 160; *n*_*controls*_ = 44,654).

The UKB is a population-based prospective study, containing a rich set of genetic and phenotypic data for approximately 500,000 participants living across the United Kingdom. Participants, aged 40 to 69 years old at recruitment between 2006 – 2010, have been linked to their annually updated electronic health records, allowing longitudinal investigation of healthcare outcomes. Similarly, AoU includes genomic data and healthcare outcomes for approximately 245,000 individuals from diverse populations in the USA.

Age distributions for UKB and AoU participants, along with delirium and all-cause dementia prevalence by age group, can be found in Supplementary Figure 1. Age distributions for the FinnGen and MGI cohort can be found in their respective original publications, that is Kurki et al. (2023; Supplementary Methods)^102^ and Zawistowski et al. (2023)^103^ respectively. Age distribution of delirium first events in FinnGen can be found here: https://r10.risteys.finregistry.fi/endpoints/F5_DELIRIUM.

### Delirium phenotype

The analysis focused on delirium episodes that were not triggered by substance intoxication or withdrawal^2^. For convenience, such delirium episodes will hereby be referred as simply delirium. Delirium cases were defined as individuals with one or more delirium-corresponding codes in their electronic health records (EHR), that is hospital inpatient, death register or primary care data. The relevant codes were: “F05” (delirium, not induced by alcohol and other psychoactive substances) for International Classification of Diseases, 10^th^ version (ICD-10)^104^ and “293.0” (Acute confusional state) for ICD-9^105^. Read v2 and v3 codes^106^ for primary care data mapping to delirium were obtained from a previously defined list by Kuan et al (2019)^107^, published in the HDRUK Phenotype Library (https://phenotypes.healthdatagateway.org/).

### Discovery of genetic risk factors

A Genome-Wide Association Study (GWAS) framework was implemented in order to identify genetic variants associated with delirium in UKB’s and AoU’s ancestrally distinct sub-populations. For this purpose, the REGENIE software (version 3.2.2) was used, which carries out a logistic regression analysis between a disease phenotype and each genetic variant, accounting for covariates, population structure and relatedness of participants^108^.

In UKB, the set of imputed genotypes was used^99^ (Data-Field 22828), filtered to include variants with > 5 minor alleles in cases and controls, imputation score > 0.5, missingness rate < 3% and deviation from Hardy-Weinberg Equilibrium (HWE) with p-value < 10^-6^. Individuals were filtered to include those with missingness rate < 5%, no mismatch between reported and genetically inferred sex (Data-Field 22001), no sex chromosome aneuploidy (Data-Field 22019), no excessive heterozygosity (Data-Field 22027) and no more than ten 3^rd^ degree relatives (Data-Field 22021). The covariates considered for the UKB GWAS included: age, sex, genotyping batch (Data-Field 22000) and the first 20 pre-computed genomic principal components (data-field 22009). Here, age was defined as age at first delirium occurrence for cases and age at last data freeze (31 October 2022) or age at death for controls. GWAS were conducted separately for sub-populations of white British ancestry (EUR; *n*_*cases*_ = 7,176 ; *n*_*controls*_ = 385,097), African (AFR; black / black British; *n*_*cases*_ = 115; *n*_*controls*_ = 7,480) and south Asian (SAS; *n*_*cases*_ = 107; *n*_*controls*_ = 9,356) ethnic backgrounds. Summary statistics from the UKB GWAS were converted from GRCh37 to GRCh38 genomic coordinates using the LiftOver software^109^. In total the analysis covered approximately 22.5, 13.6 and 8.8 million genetic variants in EUR, AFR, SAS ancestries respectively.

In AoU, short read whole genome sequencing genotypes were used^101^ for conducting GWAS on European (EUR; *n*_*cases*_ = 652 ; *n*_*controls*_ = 119,817), African/African American (AFR; *n*_*cases*_ = 233 ; *n*_*controls*_ = 52,300), and admixed American/Hispanic (AMR; *n*_*cases*_ = 117; *n*_*controls*_ = 39,977) sub-populations. The same GWAS framework as described for UKB was followed, with the exception of not including genotyping batch and principal components 11-20 as covariates, as they were not available in the AoU datasets. Sub-populations with a low number of delirium cases (<20) were excluded from the analysis. Those consisted of east Asian ethnic background in UKB and east Asian and middle Eastern genetic ancestries in AoU.

In order to increase power to detect genetic associations, our GWAS summary statistics and previously published GWAS results were combined into a multi-ancestry genome-wide meta-analysis. The METAL software (version 2020-05-05)^110^ was used to conduct a fixed effects inverse-variance meta-analysis on the set of 24,951,029 variants that were present in at least two studies. The genomic control correction method was applied in METAL to allow for multi-ancestry analysis. In total, up to 1,059,130 individuals (*n*_*cases*_ = 11,931; *n*_*controls*_ = 1,047,199) were included in the meta-analysis. Genome-wide significance was considered at a 5x10^-8^ p-value threshold. Significantly associated variants at the multi-ancestry meta-analysis were inspected for consistency at each contributing sub-cohort. Descriptive statistics for each sub-cohort are presented in Supplementary Table 1.

Wherever reported, Odds Ratios (OR) were calculated as *OR* = *e*^*β*^, where *β* is the logistic regression coefficient. 95% Confidence Intervals (CI) for the ORs were calculated as *OR*_95% *CI*_ = *e*^*β*^ ± 1.96 ∗ *SE_β_* ∗ *e^β^*.

### Sensitivity analyses

A series of sensitivity analyses were conducted in addition to our main genetic analysis, to:

- identify genetic variants associated with delirium independently of *APOE*. For this purpose, we performed a GWAS on the UKB EUR cohort conditional on the *APOE-*ε4 haplotype count (0, 1 or 2) (Supplementary Figure 2). We inferred *APOE-*ε4 haplotypes for each participant based on their rs429358 and rs7412 genotypes, as described in previous studies^35^.
- Identify genetic signals for delirium independently of dementia. To this end, we conducted in the UKB EUR cohort (a) delirium GWAS conditional on dementia (dementia-adjusted), by including all-cause dementia status as covariate (Supplementary Figure 3); (b) delirium GWAS on sub-cohorts stratified by all-cause dementia status (i.e. whether or not individuals have developed all-cause dementia at any time point) (Supplementary Figure 4); and similarly (c) delirium GWAS on sub-cohorts stratified by Alzheimer disease (AD) status (Supplementary Figure 5).
- better distinguish genetic signals in true delirium cases and controls in the elderly, by conducting a delirium GWAS on UKB EUR participants aged 60 years or older (Supplementary Figure 6).

For all sensitivity analyses, the same GWAS framework as in our main analysis of genetic risk factors was followed. All sensitivity analyses were repeated in the AoU EUR cohort for replication (significance p-value threshold < 0.05). Sample sizes for each analysis and results for rs429358, the top delirium-associated genetic variant from our main genetic risk factor analysis, are presented in Supplementary Table 2.

For the analyses that utilised dementia or AD status in UKB, the algorithmically defined all-cause outcomes were used (Data-Field 42018), provided by UKB. Briefly, disease outcomes were constructed based on self-reported medical conditions reported at the baseline assessment, ICD-9/10 codes in hospital diagnoses and procedures, and ICD-10 codes in Death Register records. The full list of ICD codes for all-cause dementia and AD is included in Supplementary Table 16 (as reported in UKBiobank Recourse 460, https://biobank.ctsu.ox.ac.uk/ukb/refer.cgi?id=460). All-cause dementia and AD diagnoses codes for the AoU cohort are reported in Supplementary Table 17.

### Multi-trait analysis between delirium and Alzheimer disease

Given the close inter-relationship between delirium and AD^6^, we wanted to quantify the shared genetic architecture between the two traits. For this purpose, we used the high-definition likelihood (HDL) software (v1.4.0)^111^ to estimate the SNP heritability (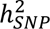) of delirium and the genetic correlation (*r_g_*) between delirium and AD using GWAS summary statistics. SNP heritability, the proportion of phenotypic variance explained by the additive SNP genetic factors^112^, was estimated in the observed scale, and converted to the liability scale using the equation: 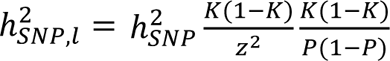, where *K* =0.015 is the population prevalence of delirium^4^, *P*=0.018 is delirium prevalence in UKB, and *z* is standard normal probability density function’s height at the quantile corresponding to cumulative probability *K*. The equation is derived from Lee et al (2011)^113^. Genetic correlation is a measure of the variance shared by two traits due to genetic causes^112^. For AD, we used the largest to-date AD GWAS meta-analysis, Bellenguez et al (2022)^58^. The AD GWAS was conducted on 487,511 European individuals (Stage I: 39,106 clinically diagnosed AD cases and 46,828 proxy AD cases) and 21 million variants. AD summary statistics were obtained from European Bioinformatics Institute GWAS Catalog (https://www.ebi.ac.uk/gwas/) under accession no. GCST90027158. Delirium summary statistics were taken from our UKB EUR GWAS sub-cohort, to better match the ancestry of the AD GWAS summary statistics and the HDL pre-computed European-ancestry reference panel.

We also applied a joint analysis of summary statistics between delirium and AD. Such approach increases statistical power to detect genetic associations for each trait^34^. The multi-trait analysis of GWAS (MTAG) software (v1.0.8) was used for this purpose^34^, jointly analysing summary statistics from our delirium meta-analysis – excluding the AoU sets – and the Bellenguez et al^58^ AD summary statistics. The MTAG analysis for the discovery of delirium genetic risk variants was conducted on 9,883,704 SNPs that overlapped across the two disorders, filtered for minor allele frequency >= 0.01 and sample size N >= (2/3) * 90^th^ percentile for each trait. MTAG results are trait-specific summary statistics (i.e., effect estimates, standard errors and p-values), interpreted similarly with single-trait GWAS results. The genome-wide significance threshold was defined as a p-value = 5x10^-8^. Independent lead variants were defined as the most significant variants within a ±500kb region, using the GWASLab python package (version 3.4.46)^114^.

The AoU EUR set was held out for replication of the MTAG lead hits. A multi-trait analysis with AD was conducted on the replication set as described above. Lead variants were considered replicated if they had a Bonferroni adjusted p-value < (0.05 / number of lead variants) and same direction of effect across the discovery and replication set.

### Mediation Analysis

We employed a mediation analysis framework in order to quantify to what extend the effect of *APOE* gene on delirium is mediated through underlying dementia status. Mediation analysis enables the decomposition of an exposure’s total causal effect on an outcome, here *APOE* and delirium respectively, into direct and indirect effect through a hypothesised mediator variable (here dementia)^53^. We used the R package ‘medflex’^53^ (v0.6-10) to calculate the so-called natural direct and natural indirect effects of the number of *APOE-*ε4 haplotypes (0,1,2) on delirium, mediated through dementia. When exponentiated, natural direct and indirect effect estimates can be interpreted as odds ratios. Medflex’s weighting-based approach was used, to estimate the unobserved potential outcomes. For the natural effect model’s parameters, bootstrap-based (n=1,000) standard errors and Wald-type two-sided p-values were obtained. Sex and age were used as covariates.

Mediation analysis, as implemented under the counterfactual framework in ‘medflex’, relies on the assumption that, after adjusting for baseline covariates, there is no unmeasured confounding between (i) exposure and outcome, (ii) exposure and mediator, and (iii) mediator and outcome^53^. It is also assumed that (iv) there are no confounders of the mediator-outcome relationship that are themselves induced by the exposure^53^. We assessed the robustness of our mediation analysis to violations of unmeasured confounding assumptions, using the E-value sensitivity analysis method^36,115^. An E-value, calculated as 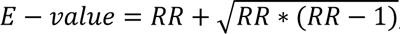, where *RR* ≈ OR, for OR > 1 is the observed direct or indirect effect risk ratio^115^, can be interpreted as the minimum unmeasured confounder effect size required to completely explain away the observed effect^36^.

We also conducted a sensitivity mediation analysis additionally adjusted for measurements of cognitive function, socioeconomic deprivation and chronic disease burden, to assess the robustness of our analysis to common dementia and delirium risk factors^4^. General cognitive ability was calculated as a composite measure – i.e. the first unrotated principal component – of four UKB cognitive tests: fluid intelligence (Data-Field 20016), numeric memory (Data-Field 4282), reaction time (Data-Field 20023, log transformed) and visual memory (Data-Field 399, log1p transformed), as described and validated in previous studies^116,117^. When cognitive tests were unavailable at baseline, measurements of the same tests at repeat assessments or online follow-up were used if available. The Townsend deprivation index immediately before joining UKB (Data-Field: 22189, decile transformed) was used as a measure of socioeconomic deprivation^118^. The Charlson Comorbidity Index (CCI) was calculated as an estimate of chronic disease burden^119^. CCI is a weighted sum of 12 chronic comorbid diseases, here calculated at the time of delirium episode. Diseases were extracted from self-reported medical conditions and cancers (Data-Fields: 20002 and 20001), and ICD-9 and ICD-10 disease codes^120^ from linked healthcare records. Weights were obtained from Quan et al^119^. This sensitivity analysis was conducted on a subset of n = 141,864 individuals, due to missing cognitive function data.

### Proteomic study population

Plasma proteome data were available in UKB for a subset of 53,075 participants. Protein measurements of 2,923 unique plasma proteins^121^ were derived from blood samples taken during randomly selected participants’ initial UKB assessment visit between 2006-2010. Proteins were measured using the antibody-based Olink Explore 3072 proximity extension assay. Proteome data have been previously undergone extensive quality control^121^. As additional filtering in the present analysis, European participants from batches 0 to 6 were extracted, as they have been reported to be highly representative of the UKB European population^121^. Moreover, protein measurements with >20% missing data were removed and the remaining proteins were mean-imputed, inverse-rank normalised and standardised (mean zero standard deviation 1) to ensure homogeneity across the proteins. Delirium incident cases were defined as the participants whose first reported delirium episode was > 1 years after baseline, that is, the date of blood sample collection at the first UKB assessment visit (Data-Field 53-0.0). Delirium data were available for up to 16 years of follow-up after baseline (censoring date: 31 October 2022). The final population consisted of 32,652 European participants and 2,919 plasma proteins. 541 participants had an incident delirium diagnosis within the follow-up period (cases). Of the remaining 32,111 participants (controls), 29,440 reached the end of follow-up without a delirium diagnosis, and 2,671 died before the end of follow-up. Follow-up information was available for all participants.

### Discovery of protein risk factors

To explore the relationships between baseline protein levels and incident delirium, we performed a proteome-wide association study (PWAS) on the UKB proteomic study population. Multivariable logistic regression models were fit between each protein as predictor and incident delirium status as outcome. In total, 2,919 models were fit, equal to the number of proteins. The models were adjusted for sex, Body Mass Index (BMI) and age at baseline. Associations were deemed significant at a Bonferroni adjusted p-value threshold: p-value < 1.7x10^-5^. For sensitivity analyses, we further adjusted protein models for *APOE-*ε4 haplotype status – zero, one or two copies of the haplotype – and for interaction between each protein and *APOE-*ε4.

We performed a pathway enrichment analysis on the set of Bonferroni-adjusted significant proteins emerging from the protein-wide association analysis. We used the Enrichr^122^ web-based tool and pathway annotations based on the Reactome^123^, Molecular Signatures Database (MSigDB)^124^ and Kyoto Encyclopedia of Genes and Genomes (KEGG)^125^ databases. The full set of Olink proteins was used as background genes. Fisher’s exacts tests were implemented to assess whether the identified proteins significantly overlap with the proteins in any of the pathways. Q-values were obtained by adjusting p-values for multiple testing using the Benjamini-Hochberg method.

### Protein selection and prediction models

To investigate whether baseline plasma proteome can improve prediction of incident delirium, a supervised machine learning approach was implemented. The LASSO (Least Absolute Shrinkage and Selection Operator) method^37^ was utilised for the selection of important proteins and to avoid overfitting given the high multicollinearity of proteomics data. For this analysis, the full set was randomly split into a training (80%; *n*_*cases*_ = 436; *n*_*controls*_ = 25,733) and test (20%; *n*_*cases*_ = 105; *n*_*controls*_ = 6,378) set. A LASSO model for binary outcomes was implemented in the training set using the glmnet R package (version 4.1.8)^126^. Here, the whole set of 2,919 proteins adjusted for demographic covariates: age, sex and BMI were used as predictors of incident delirium. In brief, the coefficients penalty parameter lambda was tuned using a 10-fold Cross-Validation (CV) framework for 100 lambdas between 10^-6^ and 0.07. The model with lambda.1se was chosen as the most parsimonious, giving the strictest model such that cross-validated error is within one standard error of the minimum^126^. To increase the robustness of the LASSO protein selection, a stability selection^38^ approach was additionally applied. For each of 100 random subsampling iterations of the training set, including all 436 delirium cases and an equal number of 436 randomly selected controls, a LASSO model as described above was fit using the penalty factor tuned in the full training set. The proteins that were selected on at least half of the subsampling iterations were chosen as robustly selected (stability-selected proteins).

Four logistic regression models with incident delirium as outcome were subsequently re-fit in the training set: (a) using only demographic covariates (age, sex and BMI) as predictors (basic model); (b) using only the stability selected proteins as predictors (proteomic model); (c) using both demographic covariates and the stability selected proteins as predictors (proteomic + basic model); and (d) using demographics, stability selected proteins and *APOE-*ε4 haplotype status as predictors (*APOE* + proteomic + basic model). For the models that included proteins as predictors (b,c and d), stepwise regression models were fit, starting with all the stability-selected proteins and removing predictors until no AIC improvement was observed.

The performance of the models was evaluated in the held-out test set. Receiver operating characteristic (ROC) curves, Area Under the ROC Curve (AUC) and Precision – Recall curves and AUC (PR-AUC) estimates were used to compare the predictive performance of the three models in the test set. Precision – Recall metrics were chosen as they are more sensitive to binary outcome imbalance^127^, as is the situation here. Two-sided Delong tests were used to compare whether AUCs were significantly different between each model pair^128^ (ΔAUC p-value < 0.05).

### pQTL analysis

We obtained protein quantitative trait loci (pQTL), that is genetic variants affecting plasma protein levels, from the UKB proteomic study population. A GWAS was conducted for each of the 2,923 inverse-rank normalised and standardised protein traits. Autosomal and X chromosome genetic variants were derived from UKB’s called genotypes (Data-Field 22418), filtered to include variants with missingness < 1%, HWE deviation p-value > 10^-15^, minor allele frequency > 1%. The proteomic study population was further filtered to include unrelated, up to 2^nd^ degree, individuals, with missingness rate < 10%. The GWAS on each protein, were conducted using REGENIE (version 3.2.2)^108^ on 30,272 individuals and ∼540,000 genetic variants. Age, age^2^, sex, sex*age, and sex*age^2^, batch, UKB assessment centre, UKB genetic array, time between blood sampling and measurement, and the first 20 genetic principal components were used as covariates.

2,464 proteins were significantly associated with at least one genetic variant (p < 5x10^-8^). Per protein, independent pQTLs were obtained through linkage disequilibrium (LD)-based clumping, including variants with LD r^2^ < 0.2 within ±250 kb windows of the top associated pQTLs. Genotype filtering and LD-clumping was performed using PLINK (v1.90b)^129^.

The independent pQTLs derived here were used as genetic instruments for protein levels in the subsequent mendelian randomisation and colocalisation analyses.

### Mendelian randomisation

Mendelian randomisation (MR) is a method used to assess potential causal influence of a modifiable exposure on an outcome, often disease risk. MR analyses are based on the use of genetic variants as instrumental variables (IV). IVs are variables associated with an exposure but not with the outcome of interest through any other pathway^130,131^. Three assumptions are required for MR to be valid: (1) IVs are significantly associated with the exposure; (2) there are no confounders of the IVs and the outcome; and (3) IVs do not affect the outcome other than through the exposure (no pleiotropy)^130^.

We assessed whether the identified protein risk factors (exposures) showed causal effects on delirium (outcome), using a two-sample MR approach^132^. Genetic instruments for protein exposure traits comprised of the per-protein independent pQTLs, as derived from our pQTL analysis in UKB (p-value < 5x10^-8^, LD r^2^ < 0.2). To further ensure no weak instrument bias in our genetic instrument pQTLs - relevant to MR assumption (1) - we also excluded pQTLs with an F-statistic < 10 in their per-protein exposure GWAS. F-statistics were approximated as 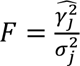 for each pQTL j, where *γ̂* is the pQTL-exposure effect size and σ its respective standard error^133^. To minimise chances of including genetic instruments with pleiotropic effects, we included only (a) pQTLs associated with less than 5 proteins (p-value < 5x10^-8^), and (b) cis-pQTLs, that is pQTLs in close proximity with their associated protein’s encoding gene. cis-pQTLs were defined as being located within ±1Mb (10^6^ base pairs) of the protein-encoding gene’s coding region. Protein-encoding genes’ start and end positions were derived as reported in the metadata from Sun et al^121^. Genetic associations of the pQTLs with delirium were derived from our UKB-EUR GWAS excluding the individuals in the proteomic set, to avoid sample overlap in our two-sample setting (*n*_*cases*_ = 6,650; *n*_*controls*_ = 354,251). The same procedure as our full UKB-EUR GWAS was followed, as described in the section ‘Discovery of genetic risk factors’.

For our main MR analysis, 1989 proteins were tested in total. The inverse variance weighted (IVW) method was used^134^ to assess causal estimates between 1,738 proteins with > 1 cis-pQTLs and delirium. For 251 proteins with 1 cis-pQTLs the Wald ratio method was used. We further employed the weighted median (WM), maximum likelihood and MR-Egger sensitivity MR methods^135^, to test for consistency of estimates with our main analysis. The MR-Egger intercept and the Cochran’s Q tests were used to assess for the presence of horizontal pleiotropy and heterogeneity in the IVs respectively^136–138^. MR associations were considered significant at an FDR q < 0.05 in our main IVW or Wald ratio MR analysis. For significant proteins, IVW or Wald ratio MR were repeated using delirium in the dementia-free UKB sub-cohort as outcome GWAS, again using an FDR q threshold of 0.05. Finally, the FinnGen delirium GWAS was used as a replication MR outcome set, following the same MR framework as described above. A significance threshold of p-value < 0.05 was used in the replication MR. Full MR results are reported in Supplementary Table 12 and 13. All MR analyses were conducted using the TwoSampleMR R package (version 0.5.11)^135^. A full list of the cis-pQTLs used, together with their association statistics with each exposure and delirium is available in Supplementary Data 3. A STROBE-MR checklist^131^ for the conducted MR analyses can be found in the Supplementary Table 18.

### Colocalisation

We tested whether genetic signals for delirium and each of the plasma proteins colocalise, i.e. the two traits are affected by the same, rather than distinct genetic variants in a specific genomic region^40^. For each of the 2,456 proteins with at least 1 pQTL, we analysed genomic regions around lead pQTLs with boundaries defined by recombination hotspot locations, based on a previously published map^139^ (https://bitbucket.org/nygcresearch/ldetect-data/src/master/EUR). We used the coloc R package (v5.2.3)^39^, specifically the Approximate Bayes Factor colocalisation method (coloc.abf), which assumes at most one causal variant per trait per region^39^. Using per-trait GWAS results and the default prior probabilities of p1 = p2 = 10^−4^ for each variant being the causal for delirium or the analysed protein respectively, and *p*12 = 10^−5^ for each variant being the causal for both, coloc.abf calculates per-variant posterior probabilities that the two traits have a common causal genetic variant in a genomic region (PP.H4 as per coloc notation). Proteins and delirium were considered to share a causal genetic variant when PP.H4 > 0.9. Suggestive colocalisation was considered for protein-delirium pairs with PP.H4 > 0.5.

### Druggability assessment

We further investigated whether selected delirium-associated proteins are suitable to act as potential drug targets. The selected proteins included those identified as significant in both our protein risk factor (p < 0.05) and MR analyses (FDR q < 0.05), as well as having a consistent direction of effect in the two analyses. We utilised the “druggable genome” resource developed by Finan et al (2017)^41^. In this resource 4,479 genes encoding drugged or druggable proteins were systematically identified, and further stratified into 3 tiers based on druggability evidence strength; as described by Finan et al: “Tier 1 (1427 genes) included efficacy targets of approved small molecules and biotherapeutic drugs as well as clinical-phase drug candidates. Tier 2 was composed of 682 genes encoding targets with known bioactive drug-like small-molecule binding partners as well as those with ≥50% identity (over ≥75% of the sequence) with approved drug targets. Tier 3 contained 2370 genes encoding secreted or extracellular proteins, proteins with more distant similarity to approved drug targets, and members of key druggable gene families not already included in tier 1 or 2.”^41^ We therefore tested whether our selected proteins overlap the with druggable genes’ encoding proteins. For proteins in Tier 1 druggability we searched the Drug Bank (https://www.drugbank.ca/) and https://clinicaltrials.gov/ databases for additional information on drug indications and clinical trial status respectively. The druggable genome can be found in the Supplementary Table 1 of the original Finan et al publication^41^.

### KGWAS

We implemented Knowledge Graph GWAS (KGWAS), a novel geometric deep learning approach that integrates GWAS summary statistics with functional genomics data (variant and gene-level annotations and interactions), to improve power in GWAS association testing^42^. Using functional genomics knowledge graphs, KGWAS estimates variants’ prior relevance to disease and recomputes the original GWAS associations in a p-value weighting framework. We used our delirium GWAMA summary statistics and KGWAS’s default fast mode, which uses Enformer embedding for variant feature and ESM embedding for gene features. We further used MAGMA^140^, as integrated in KGWAS, for gene prioritisation of KGWAS significant associations. We used a p-value threshold of 0.05 / 16,637 = 3x10^-6^ for gene-based p-values estimated by MAGMA, 16,637 being the number of genes tested.

## Acknowledgments

This work used the Edinburgh Compute and Data Facility (ECDF) (http://www.ecdf.ed.ac.uk/).

This research has been conducted using the UK Biobank Resource projects 788 and 44986. This work uses data provided by patients and collected by the NHS as part of their care and support.

UK Biobank has approval from the North West Multi-centre Research Ethics Committee (MREC) as a Research Tissue Bank (RTB) approval.

We gratefully acknowledge *All of Us* participants for their contributions, without whom this research would not have been possible. We also thank the National Institutes of Health’s *All of Us* Research Programme for making available the participant data examined in this study.

We want to acknowledge the participants and investigators of the *FinnGen* study.

The authors acknowledge the Michigan Genomics Initiative participants, Precision Health at the University of Michigan, the University of Michigan Medical School Central Biorepository, and the University of Michigan Advanced Genomics Core for providing data and specimen storage, management, processing, and distribution services, and the Center for Statistical Genetics in the Department of Biostatistics at the School of Public Health for genotype data curation, imputation, and management in support of the research reported in this publication.

This research was funded by the *Legal & General Group* (research grant to establish the independent Advanced Care Research Centre at University of Edinburgh). The funder had no role in conduct of the study, interpretation or the decision to submit for publication. The views expressed are those of the authors and not necessarily those of Legal & General.

For the purpose of open access, the author has applied a CC-BY public copyright licence to any Author Accepted Manuscript version arising from this submission.

## Data availability

Details for accessing individual-level data can be found here:

- for UK Biobank https://www.ukbiobank.ac.uk/enable-your-research/apply-for-access
- for All of Us Research Programme https://www.researchallofus.org/register

Details on obtaining delirium GWAS summary statistics used in this work can be found here:

- for FinnGen R10 release https://www.finngen.fi/en/access_results
- for MGI freeze 3 https://precisionhealth.umich.edu/our-research/michigangenomics

GWAS summary statistics generated in this work will be made publicly available via University of Edinburgh’s Datashare service [DOI to be released upon publication]. Summary statistics for the proteomic, mendelian randomisation and colocalisation analyses can be found in the Supplementary Tables file and Supplementary Data 2 and 3.

## Code availability

All software used in the present study is publicly available. The code used for all analyses in the study can be found on GitHub: https://github.com/VasiliosRaptis/deliriumGen.

