## Supplementary Figures 1-8 for "Dissecting the genetic and proteomic risk factors for delirium"

**Contents**

**Supplementary Figure 1: *Age distribution of participants in UKB and AoU.***

**Supplementary Figure 2: *Manhattan plot of delirium GWAS in UKB EUR, conditional on APOE-e4 haplotype count.***

***Supplementary Figure 3: Manhattan plot of delirium GWAS in UKB EUR, conditional on all-cause dementia status.***

***Supplementary Figure 4: Manhattan plot of delirium GWAS in UKB EUR, stratified by all-cause dementia status.***

***Supplementary Figure 5: Manhattan plot of delirium GWAS in UKB EUR, stratified by Alzheimer disease status.***

***Supplementary Figure 6: Manhattan plot of delirium GWAS in UKB EUR,* *on participants aged 60 years or older.***

***Supplementary Figure 7: Mediation analysis of APOE-ε4 on delirium, mediated by all-cause dementia, with additional covariates.***

***Supplementary Figure 8: Proteomic predictive performance on the dementia-free test set.***

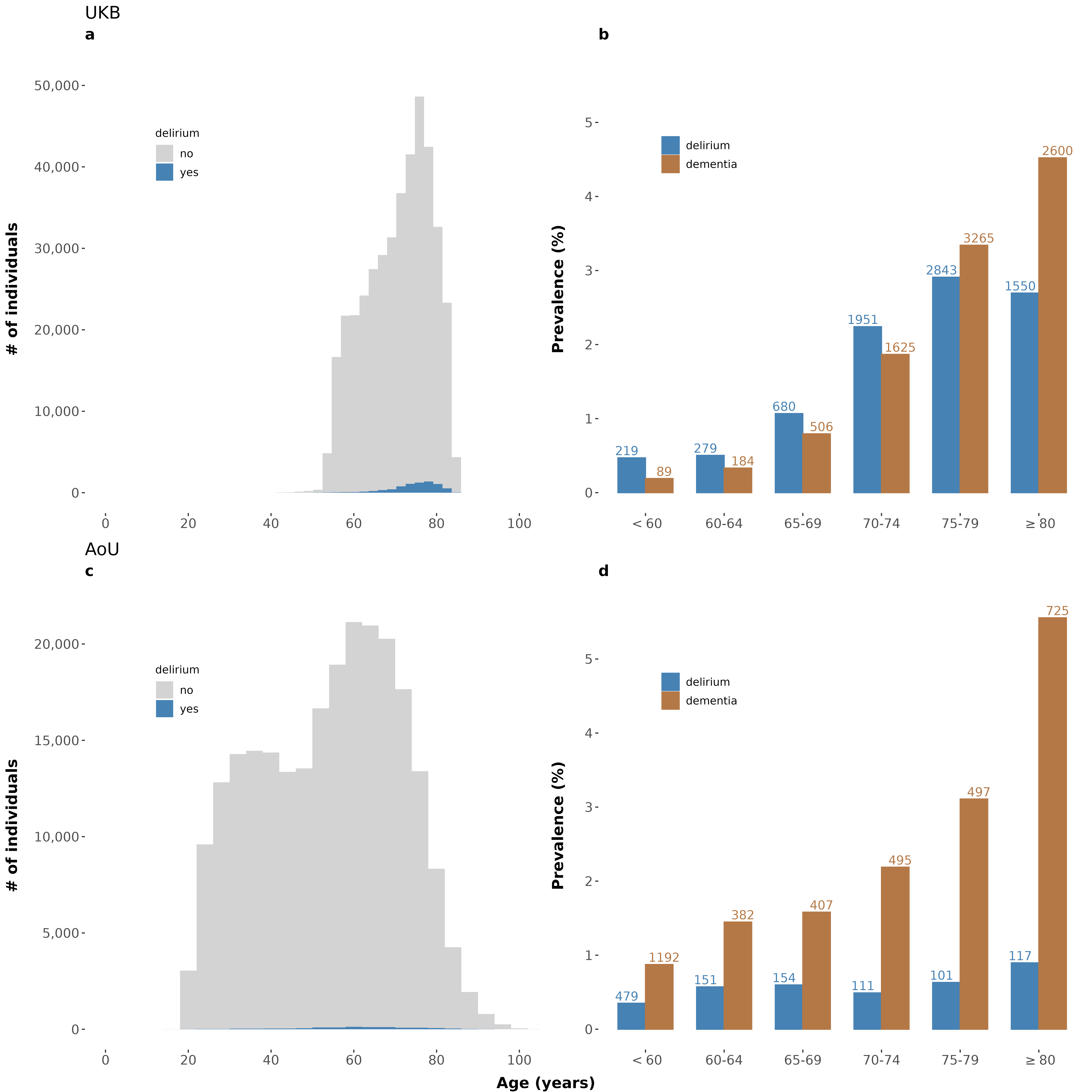

***Supplementary Figure 1: Age distribution of participants in UKB (a-b) and AoU (c-d).*** *Age of all samples used in the GWAS (n_UKB = 407,827; n_AoU = 240,158) grouped by delirium status are shown in histograms (a) and (c) for UKB and AoU respectively. Prevalence of delirium first event and all-cause dementia by age bracket is shown in (b) and (d) for UKB and AoU respectively. UKB: UK Biobank; AoU (All of Us Research Programme).*

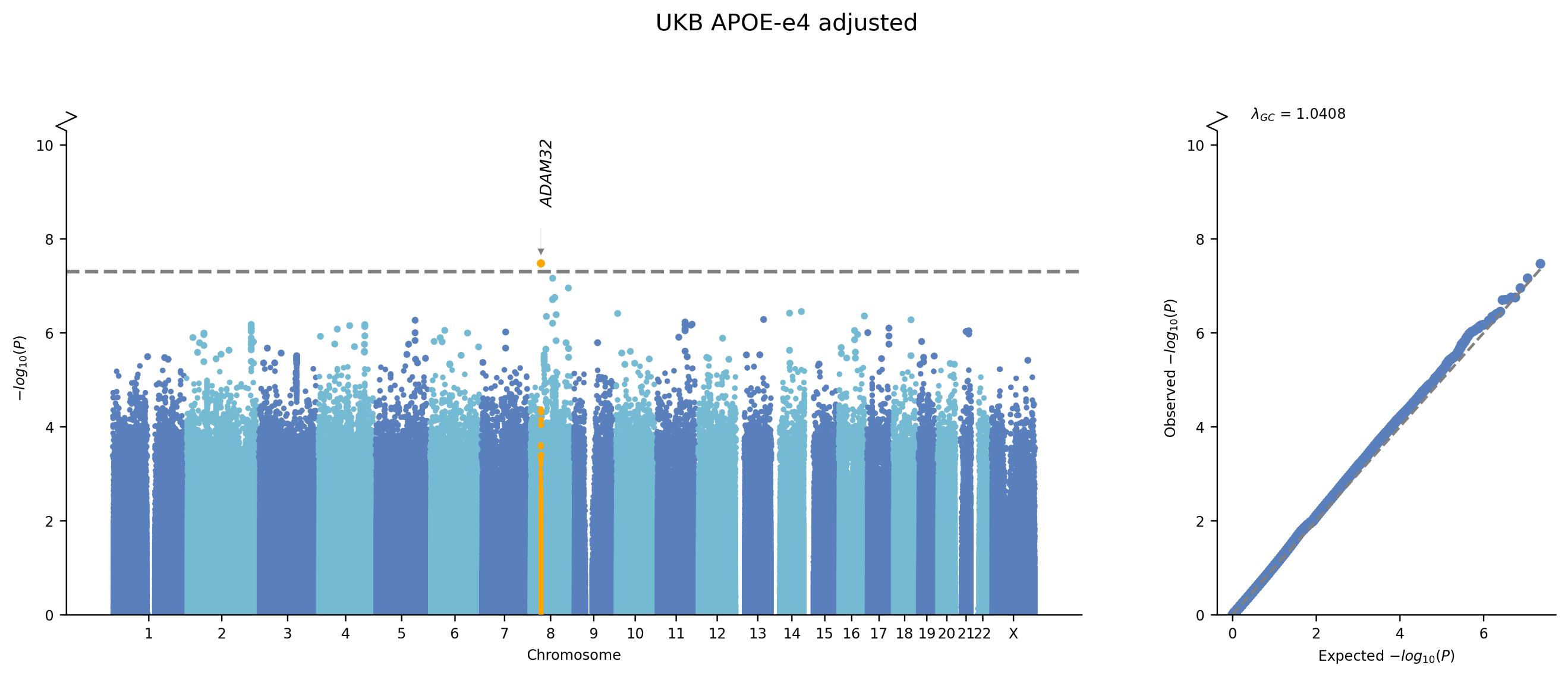

***Supplementary Figure 2: Manhattan plot of delirium GWAS in UKB EUR, conditional on APOE-e4 haplotype count.*** *(Left) Each point represents a genetic variant. The x-axis denotes the variant’s genomic position and the y-axis the GWAS association p-value. The grey dashed line denotes the genome-wide significance p-value threshold 5x10^-8^. The gene in which the lead significant variant is located is annotated. (Right) QQ-pot of GWAS observed vs. expected p-values. ADAM32: ADAM metallopeptidase domain 32; λgc: genomic inflation factor. The plot was created using the GWASLab python package.*

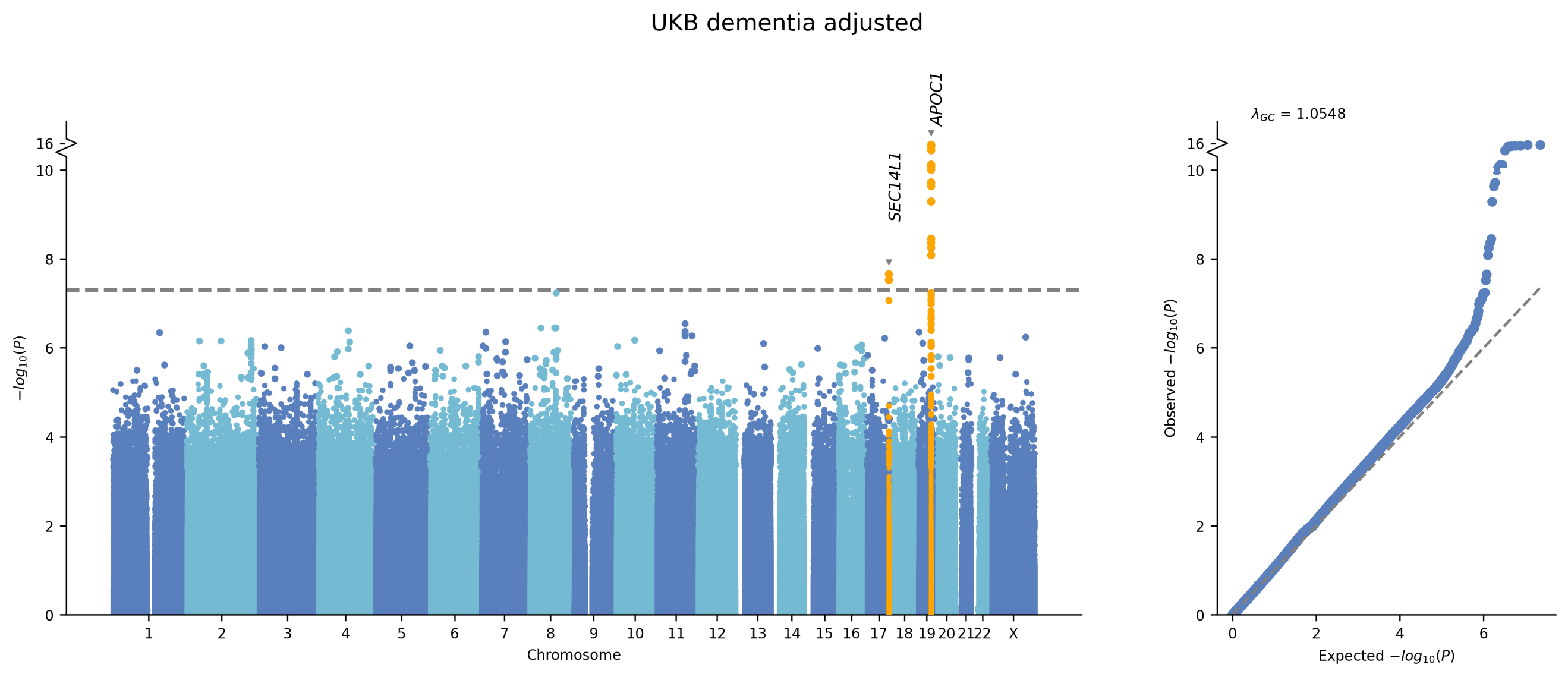

***Supplementary Figure 3: Manhattan plot of delirium GWAS in UKB EUR, conditional on all-cause dementia status.*** *(Left) Each point represents a genetic variant. The x-axis denotes the variant’s genomic position and the y-axis the GWAS association p-value. The grey dashed line denotes the genome-wide significance p-value threshold 5x10^-8^. The gene in which the lead significant variant is located is annotated. (Right) QQ-pot of GWAS observed vs. expected p-values. APOC1: apolipoprotein C1; SEC14L1: SEC14 like lipid binding 1; λgc: genomic inflation factor. The plot was created using the GWASLab python package.*

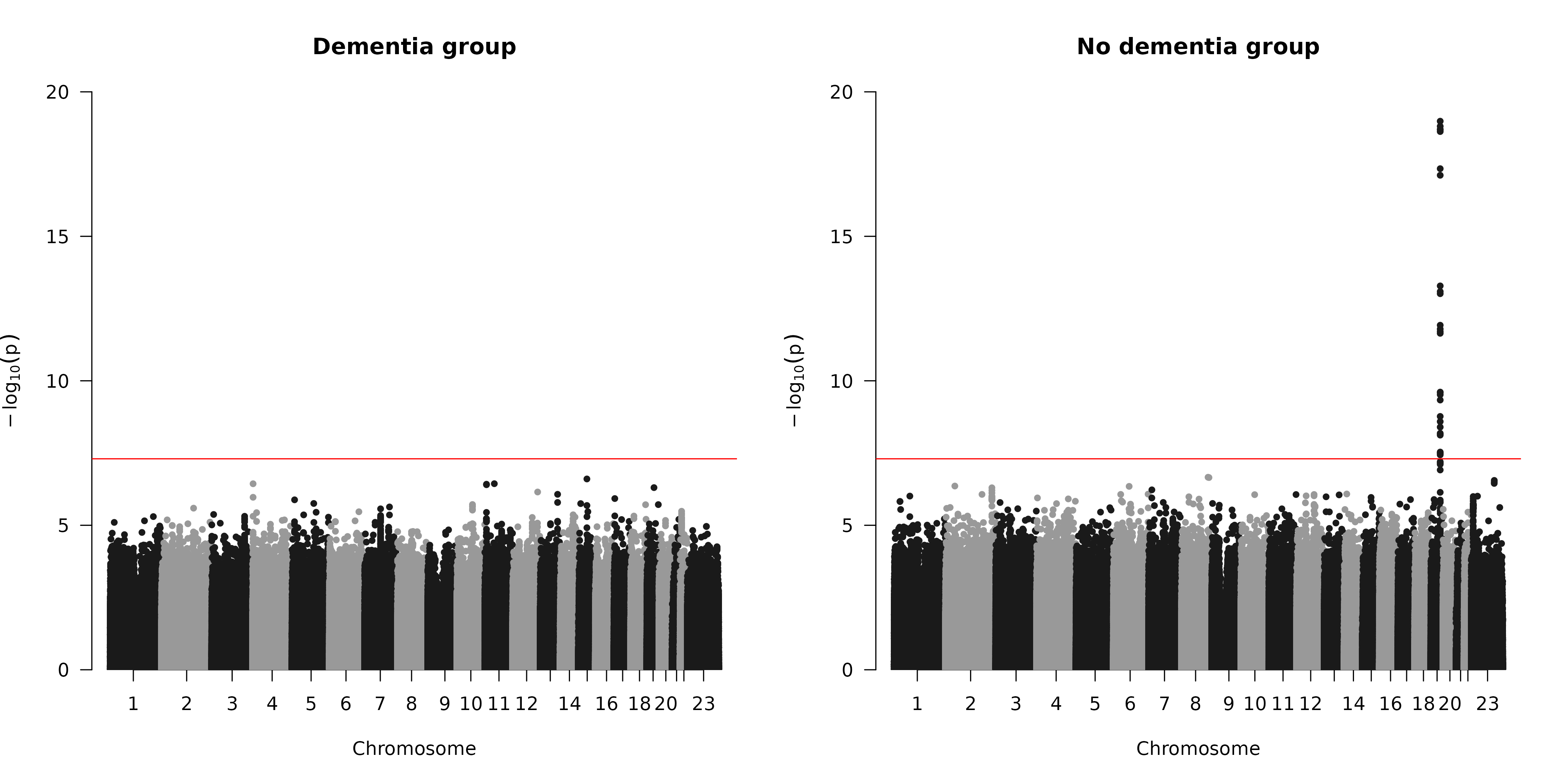

***Supplementary Figure 4: Manhattan plot of delirium GWAS in UKB EUR, stratified by all-cause dementia status.*** *Results for the dementia (n_cases = 2,561; n_controls = 5,374) (left) and non-dementia (n_cases = 4,614; n_controls = 379,642) (right) sub-cohorts. Each point represents a genetic variant. The x-axis denotes the variant’s genomic position and the y-axis the GWAS association p-value. The red dashed line denotes the genome-wide significance p-value threshold 5x10^-8^. The plot was created using the qqman R package.*

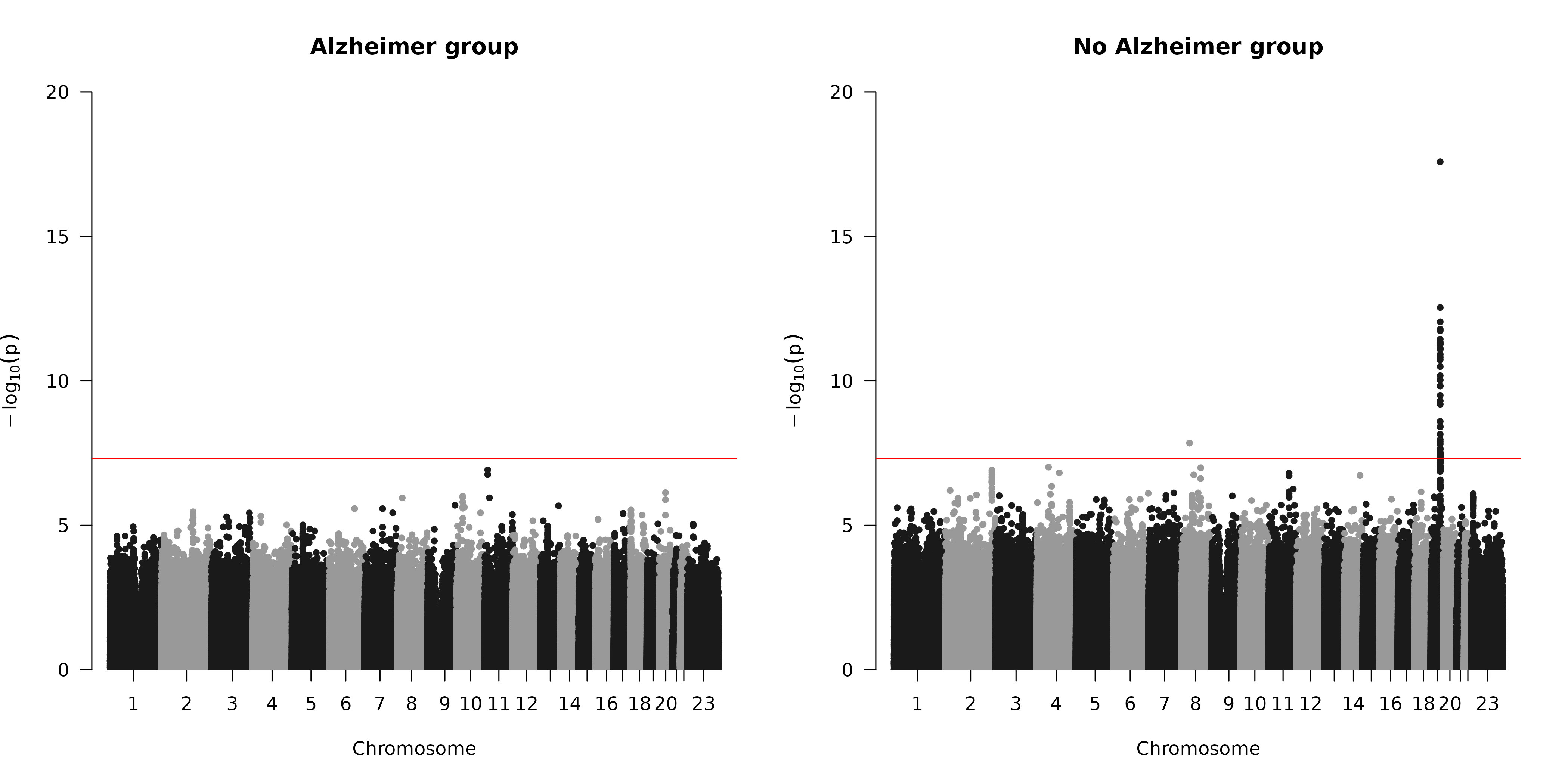

***Supplementary Figure 5: Manhattan plot of delirium GWAS in UKB EUR, stratified by Alzheimer disease status.*** *Results for the AD (n_cases = 1,142; n_controls = 2,424) (left) and non-AD (n_cases = 6,033; n_controls = 382,592) (right) sub-cohorts. Each point represents a genetic variant. The x-axis denotes the variant’s genomic position and the y-axis the GWAS association p-value. The red dashed line denotes the genome-wide significance p-value threshold 5x10^-8^. The plot was created using the qqman R package. AD: Alzheimer disease.*

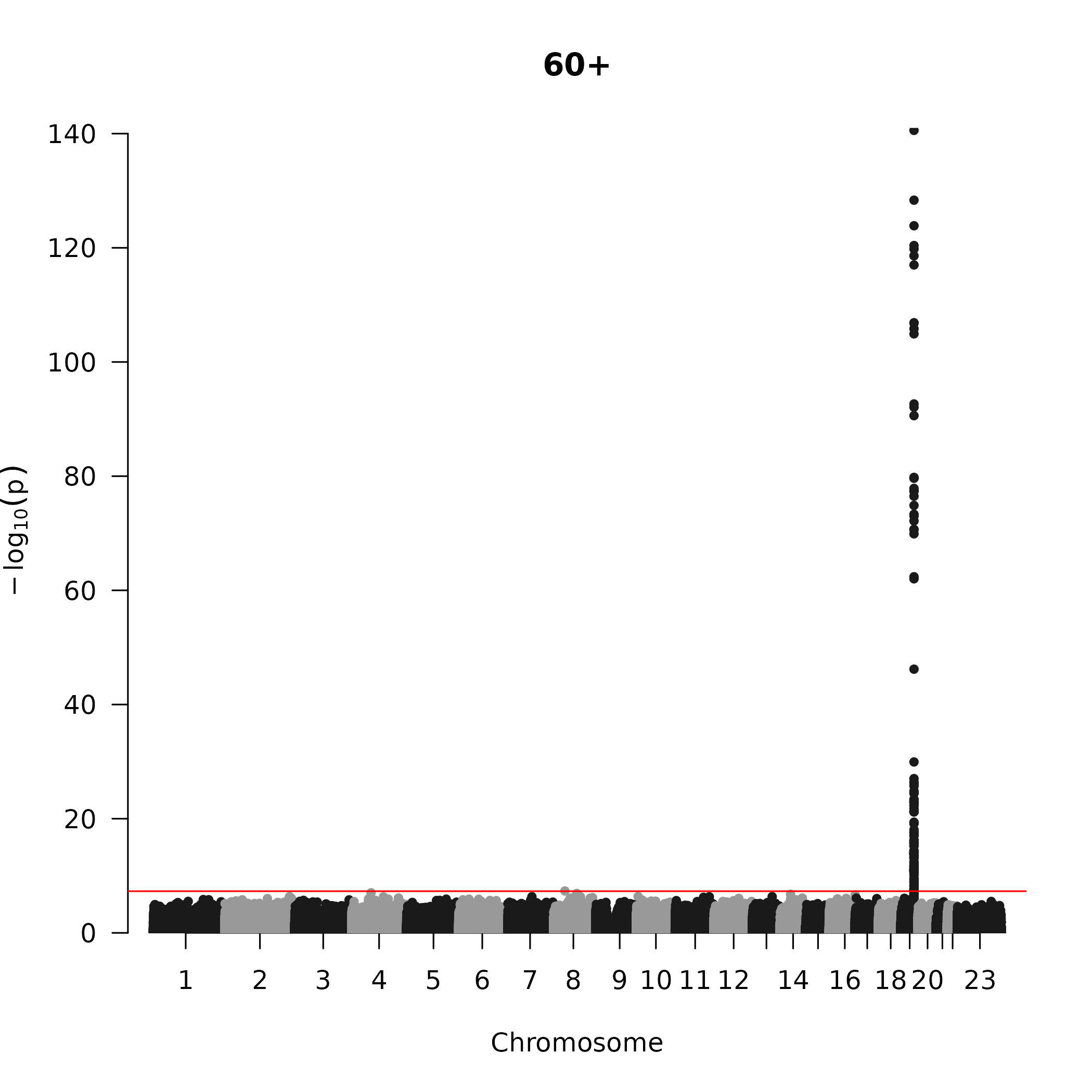

***Supplementary Figure 6: Manhattan plot of delirium GWAS in UKB EUR,*** ***on participants aged 60 years or older.*** *(n_cases = 6,955; n_controls = 335,567). Each point represents a genetic variant. The x-axis denotes the variant’s genomic position and the y-axis the GWAS association p-value. The red dashed line denotes the genome-wide significance p-value threshold 5x10^-8^. The plot was created using the qqman R package.*

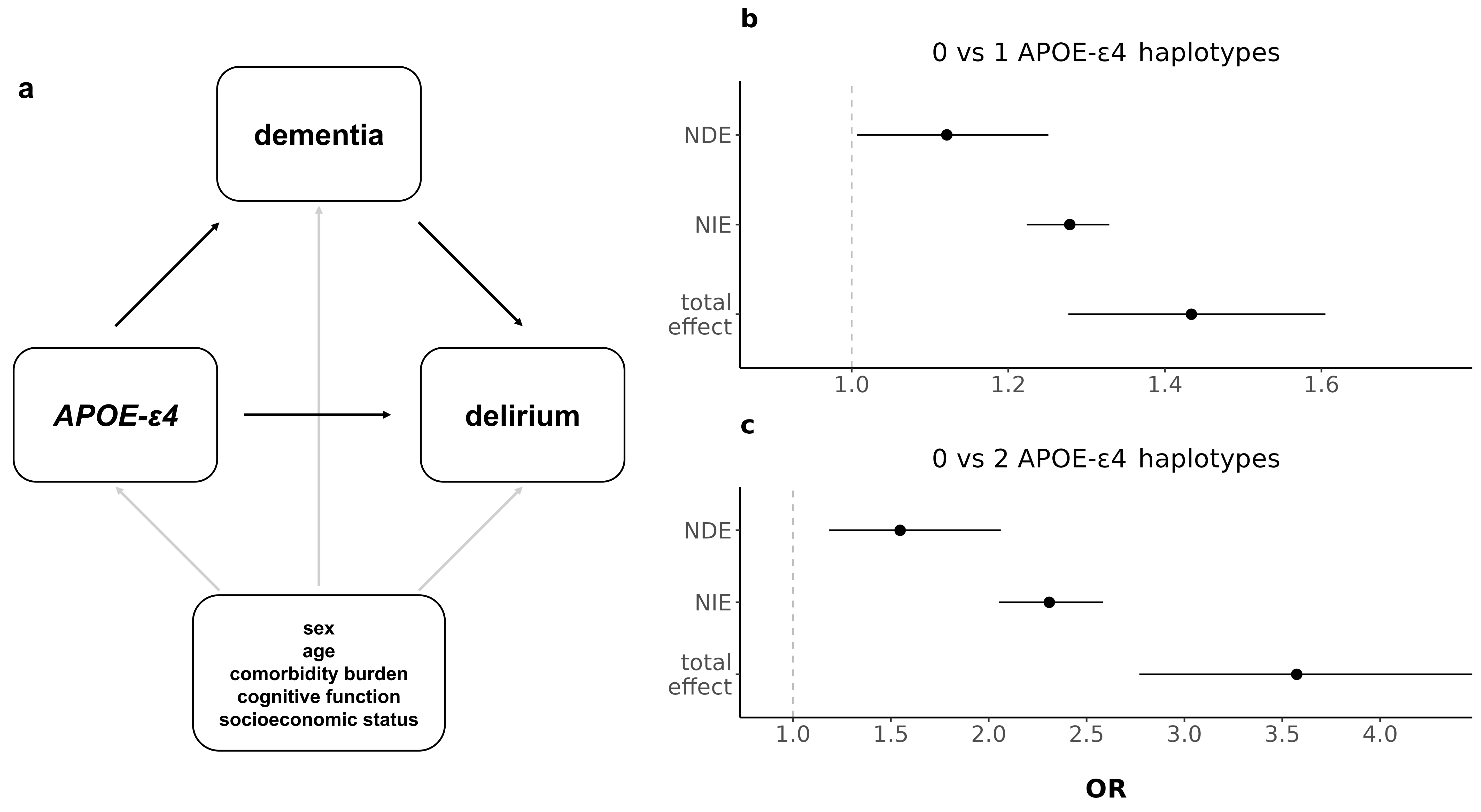

***Supplementary Figure 7: Mediation analysis of APOE-ε4 on delirium, mediated by all-cause dementia, with additional covariates. (a)*** *Hypothesised directed acyclic graph, showing all-cause dementia as mediator of APOE-*ε4 *genetic effect on delirium (black arrows), adjusted for baseline covariates: age, sex, Charlson comorbidity index, general cognitive ability and Townsend deprivation index (grey arrows).* ***(b,c)*** *Effect decomposition of the total APOE-*ε4 *effect, into natural direct (independent of dementia) and natural indirect (mediated through dementia) effects.* ***(b)*** *shows the effect of having one APOE-*ε4 *haplotype, and* ***(c)*** *the effect of having two APOE-*ε4 *haplotypes, compared to having none. OR: odds ratio. NDE: natural direct effect; NIE: natural indirect effect.*

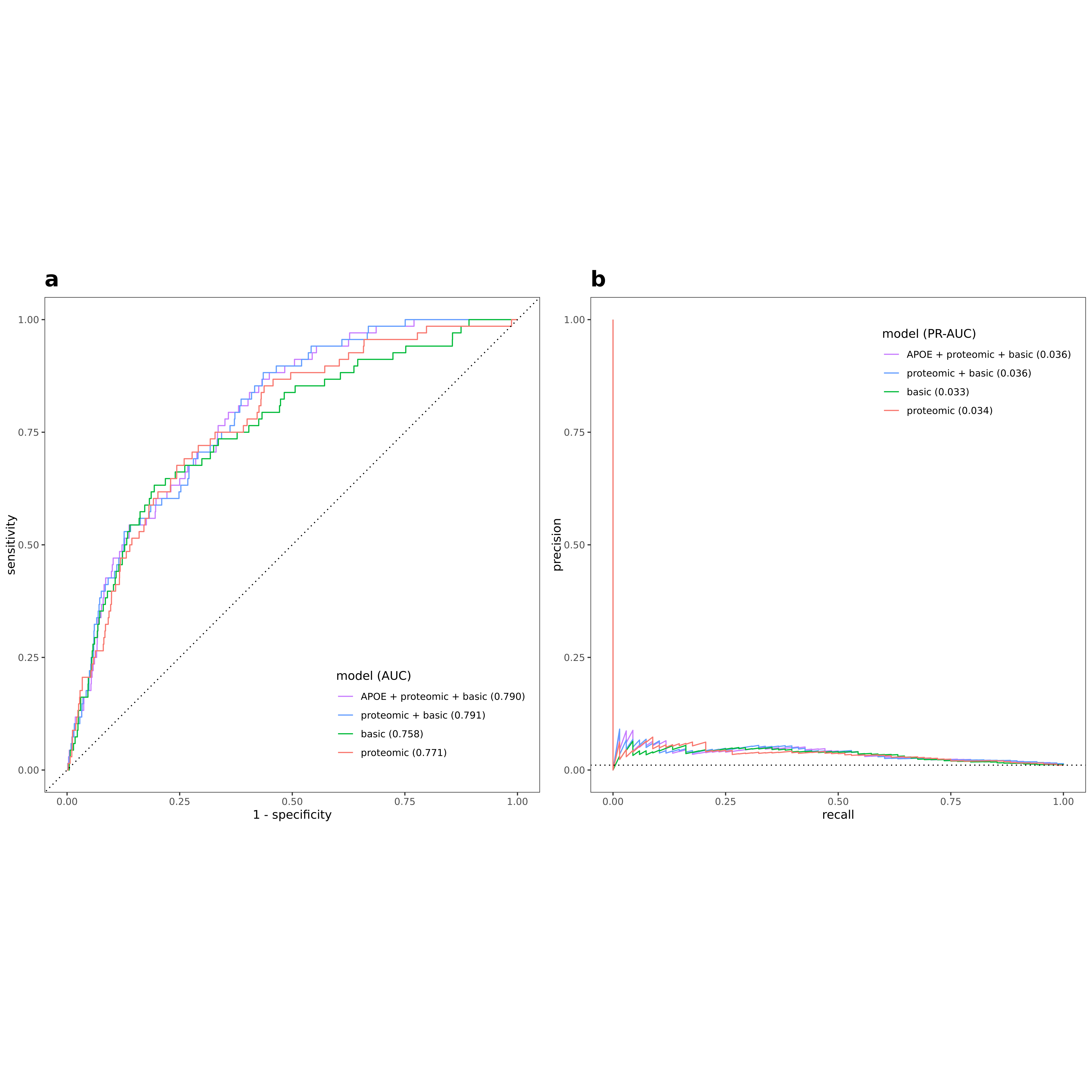

***Supplementary Figure 8: Proteomic predictive performance on the dementia-free test set.*** *Receiver Operating Characteristic (ROC) (****a****) and Precision-Recall (PR) curves (****b****) showing performance metrics of different models for predicting incident delirium on the held-out test set. The “proteomic” logistic regression model is fit with the 14 stability-selected proteins, the “basic” model is fit using only age at protein collection, sex and BMI and the “proteomic+basic” includes both. The “APOE+proteomic+basic” model additionally includes APOE-ε4 haplotype count as predictor. The dotted black lines indicate the performance of a randomly classifying model, that is the diagonal of the ROC curve (****a****) and the disease prevalence horizontal line in the PR curve (****b****). AUC: Area Under the Curve. ROC and PR plots and AUC calculations are made using the yardstick R package (v1.3.0).*
